# Impact of disease-associated chromatin accessibility QTLs across immune cell types and contexts

**DOI:** 10.1101/2024.12.05.24318552

**Authors:** Zepeng Mu, Haley E. Randolph, Raúl Aguirre-Gamboa, Ellen Ketter, Anne Dumaine, Veronica Locher, Cary Brandolino, Xuanyao Liu, Daniel E. Kaufmann, Luis B. Barreiro, Yang I. Li

## Abstract

Only a third of immune-associated loci from genome-wide association studies (GWAS) colocalize with expression quantitative trait loci (eQTLs). To learn about causal genes and mechanisms at the remaining loci, we created a unified single-cell chromatin accessibility (scATAC-seq) map in peripheral blood comprising a total of 282,424 cells from 48 individuals. Clustering and topic modeling of scATAC data identified discrete cell-types and continuous cell states, which helped reveal disease-relevant cellular contexts, and allowed mapping of genetic effects on chromatin accessibility across these contexts. We identified 37,390 chromatin accessibility QTLs (caQTL) at 10% FDR across eight cell groups and observed extensive sharing of caQTLs across immune cell contexts, finding that fewer than 20% of caQTLs are specific to a single cell type. Notably, caQTLs colocalized with ∼50% more GWAS loci compared to eQTLs, helping to nominate putative causal genes for many unexplained loci. However, most GWAS-caQTL colocalizations had no detectable downstream regulatory effects on gene expression levels in the same cell type. We find evidence that the higher rates of colocalization between caQTLs and GWAS signals reflect missing disease-relevant cellular contexts among existing eQTL studies. Thus, there remains a pressing need for identifying disease-causing cellular contexts and for mapping gene regulatory variation in these cells.

## Introduction

A major goal in complex trait genomics is to understand the biological mechanisms of trait-associated variants. To this end, a general approach has been to map molecular quantitative trait loci (molQTLs) in one or more cell types or states, and then colocalize these molQTLs with GWAS loci. GWAS loci colocalized with a molQTL are then often considered to be “explained”. To date, molQTL of gene expression levels (eQTL) have been the focus of nearly all studies. Although eQTLs have greatly improved our ability to identify genes and contexts that are impacted at many GWAS loci, over half of GWAS signals remain unexplained for most complex traits^1^.

Several groups have proposed that standard eQTL analyses from bulk samples generally identify large, but unimportant genetic effects that are shared across many cell-types^2^. Consistent with this view, an analysis of GTEx eQTLs – the largest collection of eQTLs which covers over 55 human organs – revealed that *cis*-eQTLs only mediate ∼11% of trait heritability on average^3^. These findings are often interpreted to suggest that many GWAS variants function through cell-type or context specific effects on gene regulation, motivating searches for eQTLs in specific cell-types and contexts (i.e. cell subtypes/state, disease conditions) that may be more relevant to the traits of interest. Indeed, several studies have now identified cell-type specific and highly transient genetic effects on gene expression level that colocalize with association signals at GWAS loci^4^. Even so, each study only contributes to a tiny number of additional colocalizations, raising questions as to whether this approach is effective.

More recently, two studies mapped chromatin phenotypes QTLs (cQTLs) and found that cQTLs substantially increased the fraction of GWAS loci that colocalizes with a molQTL^5,6^. For example, Aracena et al. (2024) found that chromatin accessibility QTLs (caQTLs) mediate roughly twice as much trait heritability as eQTLs and colocalize with a larger fraction of GWAS loci compared with eQTLs^5^. It is unclear why the rates of caQTLs colocalization are larger than that of eQTLs, especially given that caQTLs must impact gene expression levels in their causal path to influence human traits. Still, these findings, along with earlier reports that trait heritability explained by eQTL SNPs (11%-14%)^7^ are generally smaller than that explained by SNPs in enhancer and promoter regions (24%-79%)^8^, suggest that mapping QTLs for chromatin-level phenotypes, such as chromatin accessibility QTLs (caQTLs), may help unravel genetic mechanism of as yet unexplained GWAS loci.

To test this strategy, we built a unified map of single-cell chromatin accessibility (scATAC) profiles in peripheral blood mononuclear cells (PBMCs) from 48 individuals enrolled in three independent studies^9,10^. We integrated these data and developed novel computational approaches to map the impact of immune-related GWAS variants on chromatin accessibility across immune cell-types, contexts, and cell trajectories. Remarkably, our caQTLs colocalize with ∼50% more GWAS loci on average than bulk eQTLs, helping us to examine the genetic regulatory mechanisms that underpin previously uncolocalized immune disease-associated variants. However, by analyzing these caQTLs with immune cell-type resolved eQTLs, we find that a substantial fraction of caQTL effects do not have any eQTL effect in the corresponding cell-types or contexts. We find that these can be predicted by the lack of enhancer-promoter interactions and not by low statistical power. Thus, many genetic effects on chromatin accessibility are unlikely to be functional in most cell-types. Notably, many immune GWAS loci that colocalizes with caQTLs do not colocalize with an eQTL in any assayed cell-type, we interpret this to suggest that there are many cellular contexts in which eQTL data are missing. Thus, our findings highlight the need for expanding the catalog of eQTLs in cell-types and contexts that are more relevant to disease.

## Results

### A harmonized map of chromatin accessibility in immune cells from 59 samples

To obtain a comprehensive map of chromatin accessibility in peripheral blood mononuclear cells (PBMCs), we obtained and compared scATAC-seq data from participants with active COVID-19, COVID-19 convalescent donors, and healthy controls (**Supplementary Table 1**). We collected scATAC-seq data from the PBMCs of 25 unique donors, including 20 individuals hospitalized with COVID-19 (both in active disease and a subset at a convalescent stage) as well as 5 healthy controls. We obtained additional COVID-19 convalescent and healthy controls from two recently published PBMC scATAC-seq datasets of similar quality (i.e., in terms of TSS read enrichment and fragments per cell; **Extended Data** Fig. 1a,b). Specifically, we used 13 healthy control samples from Bengalio, et al.^9^ and 8 COVID-19 convalescent and 2 healthy control samples from You, et al^10^ (**Fig. 1a**).

**Fig. 1.**
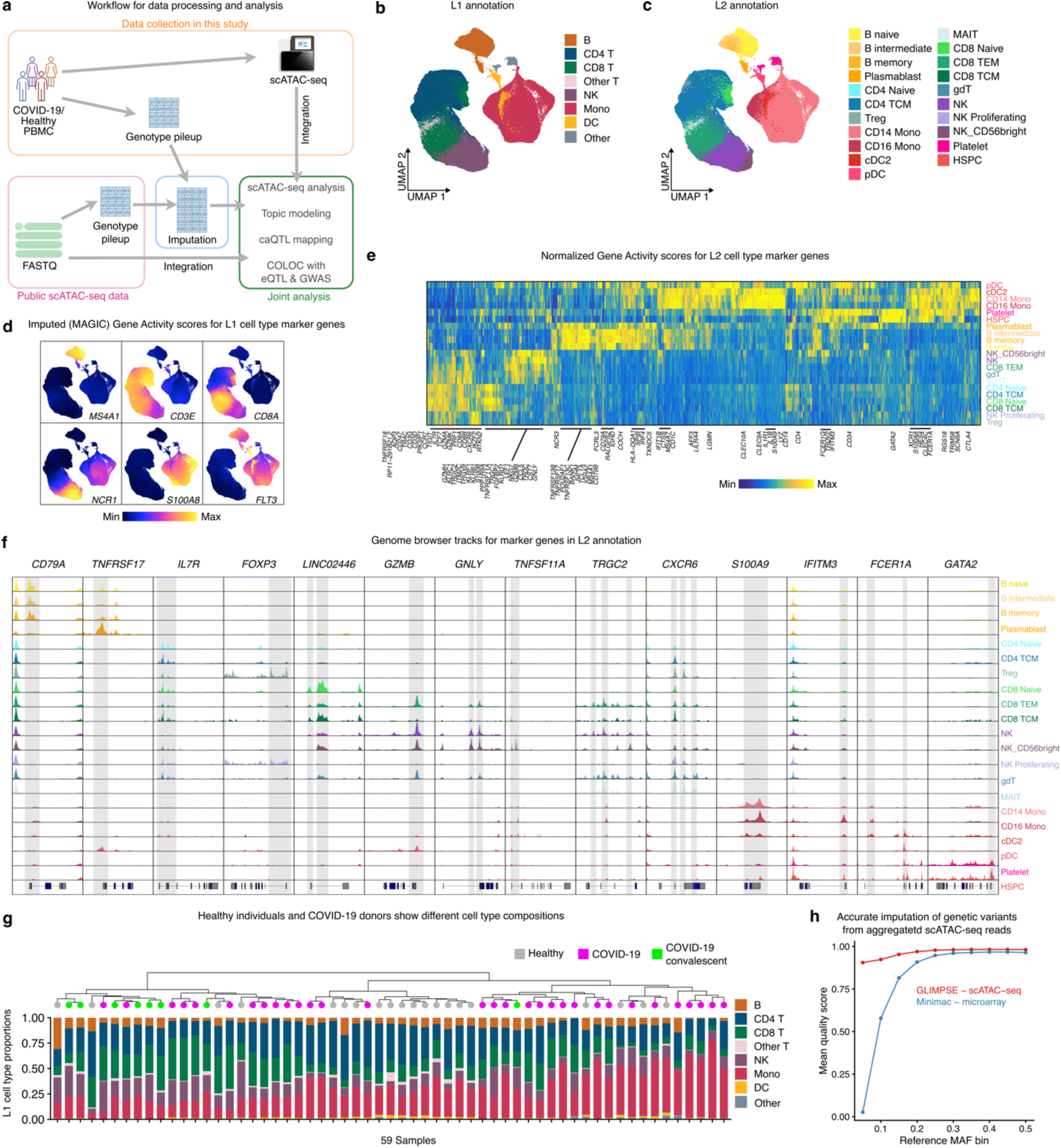
An integrated map of scATAC-seq of PBMC from 20 donors with active COVID-19, 19 COVID-19 convalescent donors and 20 healthy controls. a, Schematic representation of sample collection, data integration and analysis workflow. (Some icons are from bioicons.com). b, A UMAP embedding of all cells from three integrated studies, colored by Azimuth L1 annotation of seven common immune cell-types. c, The same UMAP embedding as in b, colored by 21 immune cell-types and subtypes in Azimuth L2 annotation. d, Gene activity scores of marker genes in the seven common immune cell-types. Scores were imputed with MAGIC for visualization purposes. e, Heatmap for marker genes for cell subtypes in L2 annotation. f, Color-coded genome browser tracks of aggregated scATAC-seq reads in genomic loci around marker genes. Shaded regions highlight cell-type-specific chromatin accessibility regions. **g**, Estimates of cell-type compositions in L1 annotation for all samples. Samples are clustered by distances in scaled cell-type compositions. **h**, Comparison of imputation quality (INFO) score from low-pass WGS using GLIMPSE, aggregated scATAC-seq reads using GLIMPSE, and DNA microarray using Minimac4 stratified by reference MAF bins.

After filtering, integration using LSI, and reducedMNN harmonization, we retained 282,424 high-quality cells for further analysis (**Supplementary Table 1**). To obtain a cell-type resolved map of chromatin accessibility, we first annotated cell-types using Azimuth^11^ from previously published COVID-19 scRNA-seq data^12^ and transferred labels to our scATAC data. We also identified 26 distinct cell clusters in this integrated dataset and confirmed that they are highly consistent with Azimuth-annotated cell-types (**Extended Data** Fig. 1c). Our final annotation has two levels of granularity: L1 annotations are later used for caQTL mapping, and contains seven major immune cell-types, while L2 annotation are used for finer-grained interpretation, and contains 21 cell-types/subtypes that are well represented in our data (**Fig. 1b-c**). We then called a unified peak set consisting of 327,746 cis-regulatory elements (peaks)^13^. To confirm the quality of these annotations, we assessed gene activity (GA) scores of marker genes in each cell-type and observed high gene activity (GA) scores at known marker genes: *MS4A1* in B cells, *CD3E* and *CD8A* in T cells, *NCR1* in NK cells, *S100A8* in monocytes, and *FLT3* in DC (**Fig. 1d**). We also visualized genome browser tracks for markers in L2 cell annotations, and found patterns of cell-type specific peaks that are broadly consistent with the annotated cell-types (**Fig. 1e-f**).

We compared the three donor groups (healthy, active COVID-19, and COVID-19 convalescent) to identify differences in cell-type compositions. While the overall cell-type compositions were similar among all individuals, there were a few notable exceptions. Two groups of COVID-19 patients had either expanded NK cell or monocyte populations, consistent with previous reports^10,14^ (**Fig. 1g**). At the L2 annotation level, we found increased proportions of memory B cells in COVID-19 patients compared to healthy controls (**Extended Data** Fig. 1d).

We next sought to obtain genotypes for scATAC-seq samples from the three harmonized datasets in order to map genetic determinants of chromatin accessibility, i.e. quantitative trait loci (caQTL). Even though one of the studies (You et al.) did not genotype individual donors, we reasoned that we could approximate low-pass whole-genome sequencing (WGS) coverage by aggregating single-cell reads from each individual and adapt GLIMPSE^15^ to accurately impute common variants in these individuals. We tested this by comparing Minimac4-imputed SNPs from genotyping arrays available from Benaglio et al. to the imputed SNPs from scATAC-seq using our GLIMPSE workflow (**Methods**). We found that genotype dosages imputed from scATAC reads and genotyping arrays were highly correlated across all reference minor-allele frequency (MAF) bins (>91%, **Fig. 1h, Extended Data** Fig. 1e-g), indicating that imputed genotypes from scATAC- reads are highly accurate and are not biased by allelic imbalance in chromatin accessibility. This allowed us to merge genotype likelihood estimates from all three studies and to perform joint imputation using GLIMPSE, resulting in a harmonized callset of 6.75 million high-quality SNPs for caQTL mapping (**Extended Data** Fig. 1h).

Altogether, we constructed a map of accessible chromatin from 282,424 PBMCs from 59 samples with high-quality, harmonized genotype information for all individuals, enabling fully-integrated downstream analysis. Imputation using aggregated scATAC-seq reads offers high-quality genotype information and our workflow (**Data availability**) can be easily adopted for future population-scale scATAC studies.

### Topic analysis of chromatin accessibility defines cell-types and states programs

Single-cell genomics data can capture gene expression in rare cell-types as well as transitional cell states. However, typical single-cell data analysis aggregates cells into discrete clusters, masking heterogeneity among cells within the same cluster. As an alternative to clustering, we applied topic modeling to our scATAC-seq data. Topic modeling represents each cell as a grade of membership (referred to as “loadings” hereafter) to inferred topics^16,17^. Each topic captures an axis of variation in the data which may represent cell-types, contexts, or biological processes. This allows us to identify peaks with differential accessibility across topics, and to measure the importance of a peak to each topic (referred to as “scores” hereafter)^18^. As such, peaks with the highest scores in each topic often reveal their associated biological functions.

We applied a topic modeling approach, fastTopics^19^, to our scATAC count matrix, and built models for six to 20 total topics (referred to as “k” hereafter, **Methods**). As expected, the number of topics greatly influences the resolution at which cell states and biological processes are captured. In our k=6 model, topics largely delineate common immune cell-types including B cells, CD4 T cells, CD8 T cells/NK cells, and monocytes. In our k=10 model, both CD8 T cells and NK cells are represented by k8, which captures the cytotoxic signatures shared between these two cell-types (**Fig. 2a**). We chose the model with 20 topics for all downstream analyses, as it captures the majority of common cell-types and recognizable cell states in our PBMC data (**Fig. 2a**, **Extended Data** Fig. 2).

**Fig. 2.**
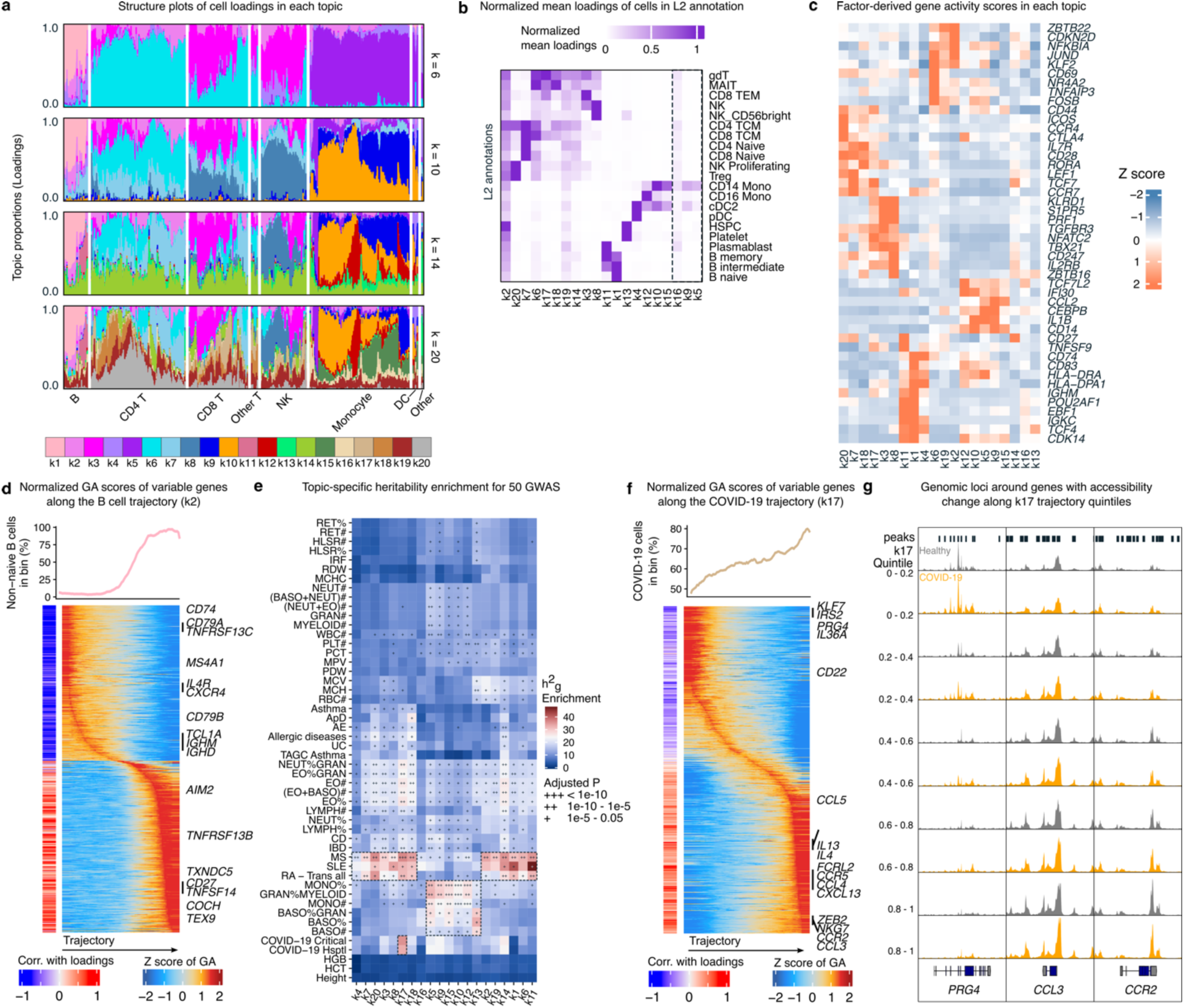
Topic modeling helps interpretation of inter-cellular and inter-individual variation in scATAC-seq profiles. **a**, Structure plots of topic loadings in 2,000 randomly selected cells when fitting 6, 10, 14 and 20 topics. Cells were grouped by seven common immune cell-types to highlight coarse-grained differences in topic loading among cell-types. **b**, Heatmap showing the average loading for each topic in each cell-type in L2 annotation. **c**, Heatmap of Z-score normalized gene-level scores calculated from peak-level scores in each topic. **d**, Top, smoothed percentage of non-naive B cells in trajectory percentile. Heatmap on the left shows the Spearman correlation between gene-activity (GA) scores and memory B cell trajectory; heatmap on the right shows row-normalized GA score changing along the trajectory. **e**, Heritability enrichment of 50 GWAS using peaks with the highest 10% of scores in each topic. Dashed boxes highlight specific enrichment results discussed in the main text. **f**, Similar to **d**, showing the trajectory and relevant genes in COVID19-associated topic k17. **g**, Genome browser tracks of the genomic region around three genes (*PRG4*, *CCL3* and *CCR2*) that progressively gained or lost accessibility along k17 trajectory. Cells are grouped by disease status and k17 quintiles.

We observed that multiple topics were almost exclusively used by certain cell-types, including B cells (k1, k11), monocytes (k10, k12), and DC (k4) (**Fig. 2b**). In contrast, T cell subsets often map to more than one topic, highlighting the subtle differences across T cell subsets and cell states, and the challenges in using clusters to represent T cell states. One topic, k2, was ubiquitously present in all cell-types. Upon investigation, we found that k2 loadings are highly correlated with the TSS enrichment score. The top 3,000 peaks with the highest scores in k2 are over-represented in promoter regions (p-value < 2e-16, hypergeometric test; **Extended Data** Fig. 3a,b), suggesting that k2 likely represents single-cell data quality rather than biological variation.

To functionally annotate the different topics, we derived gene-level scores in each topic from the peak-level scores. We tested four different peak-to-gene mapping strategies and found that distance-based exponential weighting function from ArchR performed the best on our benchmarking (**Extended Data** Fig. 3c**, Methods**)^20^. Using this strategy, we identified a set of genes driving each topic and we observed well-known cell-type markers among the highest-scoring genes, including *EBF1 and CD83* for naive B cells (k1); *CD27* and *TNFSF9* for memory B cells (k11); *CD247* and *ZBTB16* for NK cells (k8); *S1PR5*, *KLRD1*, *PRF1* and *TBX21* for cytotoxic CD8 T cells (k3); and *ICOS* and *CTLA4* for Treg cells (k20) (**Fig. 2c**, **Supplementary Table 2**). The high-scoring genes in each topic were also enriched in relevant biological process gene sets (**Extended Data** Fig. 3d; **Supplementary Table 3**). Finally, we tested the enrichment of transcription factor (TF) binding motifs in the top 3,000 peaks with the highest scores for each topic (**Supplementary Table 4**). Again, TFs known for immune cell-types and states show significant enrichment in the corresponding topics (**Extended Data** Fig. 3e).

Cell transitions from one state to another (e.g. during disease progression) generally exhibit a continuous rather than a discrete change in gene expression^21,22^. We reasoned that the loading of topics corresponding to a cell state transition captures the cell trajectory along possible transitions. Indeed, we identified evidence that topic k1 loadings represent the transition between naive B cells to memory B cells (**Methods**). For example, we observed progressive enrichment of non-naive B cells (including memory B cells and plasmablasts) along the k1 trajectory (**Fig. 2d**, top). We also adapted the ArchR getTrajectory algorithm to identify genes with changing activities along memory B cell trajectory and observed decreasing GA scores for naive B cell marker genes (*IL4R*, *TCL1A*, *IGHM*, *IGHD*) and increasing GA scores for memory B cells marker genes (*AIM2*, *CD27*, *COCH*) (**Fig. 2d**, bottom) (**Supplementary Table 5**). Thus, cell loadings from our topic modeling effectively and directly capture biologically-meaningful trajectories along cell states.

Finally, we determined the relevance of each topic in terms of explaining complex disease heritability. We used stratified LD score regression (s-LDSC)^8^ to calculate heritability enrichment of each topic across 50 GWAS traits (**Supplementary Table 6**). As expected, we observed greater h^2^g enrichment in many immune-related diseases and blood phenotypes compared to height, a trait used as negative control. Notably, we found large h^2^g enrichment for three autoimmune diseases (RA, systemic lupus erythematosus [SLE] and multiple sclerosis [MS]) in the lymphoid-related topics (**Fig. 2e**). For SLE, B cell topics (k1 and k11) are the most enriched, consistent with the known role of B cells in SLE etiology^23,24^. In addition, monocyte and myeloid-related GWAS (MONO%, MONO#, GRAN%MYELOID) are enriched in monocyte-related topics (k5, k9, k10, k12, k13, k15; **Fig. 2e**), in line with the expectation that cellular programs in monocytes causally regulate myeloid cell numbers and proportions. Interestingly, we found that one topic, k17–which was found in some CD8 T and non-classical T cells–was significantly enriched for heritability for both hospitalized and critical COVID-19 GWAS (**Fig. 2e**).

Thus, our analyses identify cell states and regulatory programs that likely mediate genetic risks for several complex immune traits.

### Topic-derived cell trajectories identify COVID-19–associated cell state continuums

We next sought to find topics that may be associated with COVID-19. As we found k17 to be significantly enriched for COVID-19 GWAS heritability, we tested the association between k17 loadings and cell donor COVID-19 status to assess whether cell donor active COVID-19 status explains a significant proportion of variation in loadings (**Methods**). Indeed, k17 is significantly enriched for COVID-19 cells (p-value = 0.023). Topic k17 largely represents CD8 TEM, and cells with high k17 loadings are disproportionately from COVID-19 donors (**Fig. 2f**, top), suggesting an expanded CD8 TEM population in COVID-19 patients.

To further investigate k17, we built a trajectory using k17 loadings and grouped cells by trajectory quintiles (**Fig. 2f**). We found that several genes previously linked to COVID-19 showed varying chromatin accessibility along the k17 trajectory (**Supplementary Table 7**). For example, we observed decreased accessibility around the *PRG4* gene body along the k17 trajectory^25^, while accessibility increased around *CCL3* and *CCR2*^26^ (**Fig. 2g**). Taken together with the independent observation that high-scoring k17 peaks are enriched in COVID-19 heritability (**Fig. 2d**), our results suggest that an expanded CD8 TEM cell population in peripheral blood is a hallmark of COVID-19 and that CD8 TEM cells–or related cells–are important mediators of COVID-19 genetic risk.

More generally, we compared GA scores in COVID-19 cells across quintiles against healthy cells. COVID-19 cells in higher quintiles exhibited more differentially active genes than those in lower quintiles (**Extended Data** Fig. 3f, **Supplementary Table 8**). Notably, these differentially active genes were enriched in pathways related to chemotaxis, immune cell migration and T cell activation, consistent with the immune response upon COVID-19 infection (**Extended Data** Fig. 3g). These findings complement our observation that k17 is enriched with COVID-19 cells and COVID-19 heritability and could not be replicated using a cluster-based approach. These results demonstrate the utility of modeling continuous cell states using a topic modeling framework.

### A high-resolution map of caQTLs in PBMCs

We next used our harmonized dataset to map the impact of genetic variation on chromatin accessibility in multiple cell-types. To map caQTLs, we first used RASQUAL to model both intra-individual allelic-imbalance and inter-individual variation in chromatin accessibility for SNPs in a 10 Kb window flanking the peak center in PBMCs (aggregating all cells from a donor) as well as the seven immune cell-types defined in our L1 annotation. We used phenotypic principal components (PCs), genotype PCs, and multiple quality control measurements^27^ as covariates (**Methods**, **Extended Data** Fig. 4a). In total, we identified 37,390 caQTLs (corresponding peaks are referred as cPeaks hereafter, making up 11.7% of all tested peaks), 8,792 of which were discovered in L1 cell-types (**Fig. 3a**), but not when aggregating all PBMC cells. This far surpasses the number of significant caQTLs identified in previous studies^9,28^.

**Fig. 3.**
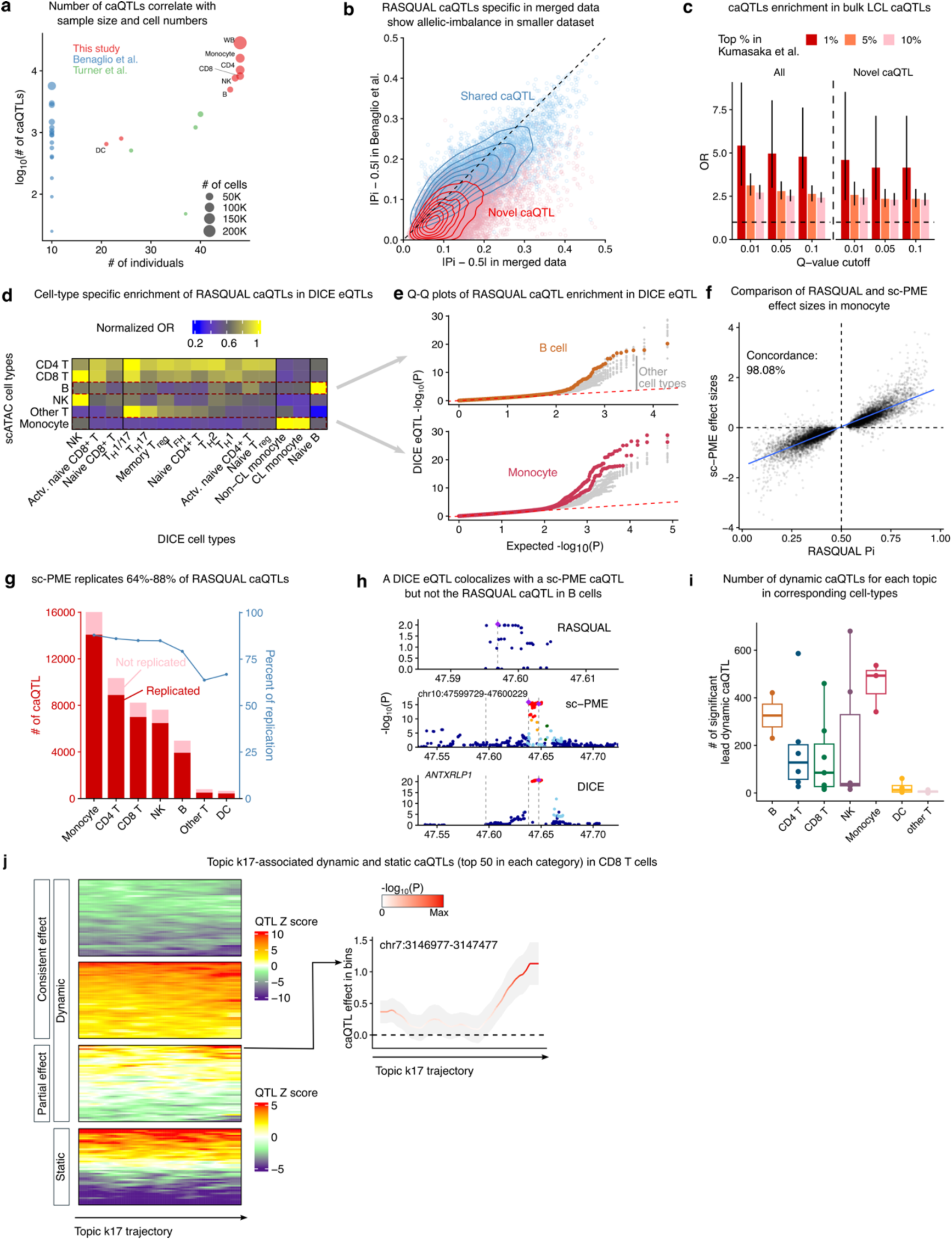
Mapping of caQTL with RASQUAL and sc-PME model. **a**, Scatter plot comparing the number of RASQUAL caQTL as a function of cell number and sample size in this study and two other published scATAC-seq caQTL studies. **b**, RASQUAL effect sizes in our merged data and Benaglio et al. data are highly correlated. Novel caQTLs found in merged data show allelic imbalance in Benaglio et al., albeit with smaller effect sizes. **c**, RASQUAL caQTLs in whole blood are enriched in bulk caQTLs from LCL for all significant ones (left) and only novel ones found in our merged data (right). **d**, Heatmap showing cell-type-specific enrichment of caQTLs in DICE eQTLs. Odd ratios are normalized by the maximum value in each row. **e**, QQ-plot of RASQUAL caQTLs in DICE eQTLs. Top: caQTLs in B cells show elevated signals only in eQTLs from B cells in DICE; bottom: caQTLs in monocytes show elevated signals only in eQTLs from classical and non-classical monocytes in DICE. All other DICE cell-types were colored grey. **f**, RASQUAL and sc-PME caQTLs have highly correlated and concordance effect sizes. Results from monocytes are used as an example here. **g**, Replication of RASQUAL caQTL in sc-PME model in seven common immune cell-types. Barplot shows the number of RASQUAL caQTLs that are replicated or not in sc-PME model; line chart shows the percentage of RASQUAL caQTLs replicated. **h**, An example of a DICE eQTL to gene *ANTXRLP1* in monocytes colocalizing with sc-PME caQTL but is different from the RASQUAL lead SNP. Vertical dashed lines highlight the genomic coordinations of lead SNPs in RASQUAL, sc-PME and DICE. The shaded region highlights the mapping window of RASQUAL and its position relative to the mapping window of sc-PME. SNPs are colored by LD to the lead SNP. **i**, Number of dynamic caQTLs in each cell-type along relevant trajectories defined by topic loadings. **j**, left: Z scores of dynamic and non-dynamic caQTLs from CD8 T cells in rolling windows along topic k17 trajectory. Dynamic caQTLs were further categorized by whether they are consistently active along k17 trajectory or only in part of the trajectory. The most significant 50 caQTLs from each category were plotted. Right: effect sizes of one representative dynamic caQTL.

To verify that the caQTLs we found are likely true positives, we separately mapped PBMC caQTLs in each of the three scATAC-seq datasets. We found multiple lines of evidence indicating that the caQTLs uniquely identified in our harmonized data (“novel caQTLs”) are *bona fide* caQTLs. First, novel caQTLs showed allelic imbalance in the smaller dataset from Benaglio et al., despite being non-significant (**Fig. 3b**). Second, both our total caQTL set and novel caQTLs are strongly enriched in bulk caQTLs from lymphoblastoid cell lines (LCLs) (**Fig. 3c**)^29^. Finally, caQTLs in L1 cell-types showed cell-type specific enrichment in eQTLs from 15 immune cell-types in the DICE consortium^1,30^. For instance, caQTLs in monocytes and B cells are the most enriched for eQTLs in classical/non-classical monocytes and naive B cells in DICE, respectively (**Fig. 3d**). Similarly, caQTLs in CD4 T cells were broadly enriched in eQTLs across various T cell subtypes in DICE. When visualizing the p-value distribution of caQTL SNPs in DICE eQTLs, a similar trend emerged (**Fig. 3e**). We also obtained single-cell eQTLs identified from scRNA-seq data collected from 63 COVID-19 and 106 control donors, of which 25 donors overlap ours (**Randolph et al.).** Again, we found cell-type-specific enrichment of caQTLs in eQTLs in matched cell types (**Extended Data** Fig. 4b). Together, this evidence suggests that our caQTLs are likely to be true positives.

The goal of our work is to determine the extent to which caQTLs can explain immune disease-associated loci. However, RASQUAL does not report effect size and standard error of identified caQTLs, which complicates statistical integration of RASQUAL caQTLs with GWAS summary statistics. Additionally, caQTLs mapped using RASQUAL are biased toward heterozygous SNPs with high read coverage, as SNPs outside peaks have weaker or no allele-specific signal, and lower phasing accuracy. To adapt single-cell caQTL data for colocalization analysis^31^, TWAS^32,33^, mashr^34^ and meta-analysis^35^, we used a single-cell Poisson mixed-effects model (sc-PME)^36^ to generate standard summary statistics of caQTL effects. In addition to providing critical statistics for downstream statistical analyses, sc-PME allows a larger mapping window (250Kb) compared to the 10Kb used in RASQUAL. This window is difficult to extend in RASQUAL because it requires haplotype phasing, which worsens as a function of distance. Yet, a larger mapping window may help capture additional significant caQTLs (**Extended Data** Fig. 4c).

To reduce computational time and avoid inflated p-values due to high dropout rates in scATAC count data, we restricted our analysis to the 37,390 significant cPeaks from RASQUAL in PBMC and L1 cell-types. As expected, we did not find many caQTLs in underrepresented cell-types (5,006 caQTLs in DC and 3,662 caQTLs in “Other T Cells”, 10% FDR). However, in the other 5 immune cell-types, we identified 10,739-22,785 caQTLs (mean: 16,297; 10% FDR). We then compared sc-PME results to those from RASQUAL, focusing specifically on the effect size estimates at top caQTL SNPs. We observed high concordance between the two approaches (e.g. 98.08% in monocytes; **Fig. 3f**), supporting the validity of the sc-PME results. We also found that across L1 cell-types, an average of 79.1% of RASQUAL caQTLs are replicated by sc-PME. As anticipated, the rate of replication is lowest among rare cell-types (**Fig. 3g**). Finally, compared to a single-cell linear mixed-effects model, effect sizes from sc-PME had higher reproducibility and correlation with RASQUAL caQTLs (**Extended Data** Fig. 4d).

We hypothesized that sc-PME would allow us to map caQTLs missed by RASQUAL owing to larger testing window sizes. To test this, we analyzed 1,337 peaks in B cells whose sc-PME lead QTLs are more than 10 Kb away from the cPeak center but reside in distal chromatin accessibility peaks, as these cases are more likely to capture distal causal effects. These distal peaks are enriched in cPeaks (8.9% versus 3.4% genome-wide, p<2e-16; hypergeometric test), suggesting that sc-PME captures *bona fide* distal caQTLs that are missed by RASQUAL. We highlight a peak (chr10:47599729-47600229) whose lead RASQUAL caQTL is located within the cPeak itself, but the lead sc-PME caQTL is ∼50 Kb upstream, is much more statistically significant, and colocalizes with an eQTL for gene *ANTXRLP1* in B cells (PP4=0.95, **Fig. 3h**). Thus, beyond providing critical summary statistics (i.e. Z-scores) that can be used for downstream statistical analyses, sc-PME can also help capture causal SNPs that are invisible to RASQUAL.

In addition to identifying caQTLs in the L1 cell-types, we identified dynamic genetic effects along cell trajectories as defined by loadings from our topic modeling. Specifically, we tested for the linear interaction between lead caQTL in the L1 cell-types and the loadings of each topic (**Methods**), which capture a continuum of cell states along different axes of biological variation. We only mapped dynamic caQTLs in each cell type to avoid confounding with cell-type specific caQTLs. In total, we identified 4,200 peaks (Q-value < 0.01) that have at least one dynamic caQTL in one or more cell-type-topic pairs. On average, we detected 158 significant dynamic caQTL in each cell-type-topic pair and, as expected, we found that the number of cells in a topic greatly influenced the statistical power for calling dynamic caQTLs (**Fig. 3i**).

To showcase examples of dynamic caQTLs, we highlight k17-interacting caQTLs in CD8 T cells. We divided dynamic caQTLs into two groups: (i) Consistently significant, which are significant along the entire k17 trajectory, but with varying effect sizes, and (ii) Partially significant caQTLs, which have significant effects in parts of the trajectory. The variability in the effect sizes of these caQTLs along the k17 trajectory is visualized and contrasted with that of randomly chosen non-dynamic caQTLs (likelihood-ratio test p-value > 0.5) (**Fig. 3j**). To map dynamic caQTL peaks to genes, we used the Activity-by-Contact (ABC) model and found that genes associated with k17 dynamic caQTLs were enriched in immune and disease-related pathways, including natural killer cell mediated cytotoxicity (GO:0002228, p-value=2.61e-8) and regulation of lymphocyte activation (GO:0051249, p-value=3.36e-7) (**Extended Data** Fig. 4e). Additionally, several linked genes in ABC model were also implicated in the k17-associated COVID-19 genes we identified in topic analysis, including *IFNLR1, IL5RA, IL6ST, KLRC4, KLRD1, KLRK1, SYNGR1, TLR1* and *TNFSF14*.

In summary, we established a map of 37,390 static and 4,200 dynamic caQTLs in PBMC and common immune cell-types. Not only do these caQTLs capture the impact of genetic variants on chromatin accessibility in common immune cell-types, but they also capture dynamic effects that manifest in important cell contexts such as cytotoxic cells. Summary statistics for all caQTLs can be readily used in standard and popular downstream analyses and are publicly available (Zenodo: TBD).

### Sharing and specificity of caQTLs and eQTLs across cell-types and states

Observations from single cell RNA-seq studies suggest that eQTLs and caQTLs are largely cell-type-specific^9,28,37^. In contrast, molQTL maps derived from bulk RNA-seq data suggest that most QTL effects are shared across cell-types^1,38–40^. We sought to systematically evaluate the specificity of caQTL effects across the seven immune cell-types in our L1 annotation, and to evaluate their cell-type specific impact on gene expression levels.

We first compared the sharing of our sc-PME caQTLs. The general picture that emerged is that the majority of caQTLs are shared among all seven immune cell-types, except for monocytes, for which 42.5% of caQTLs identified were monocyte-specific. Still, only an average of 18.8% of caQTLs were cell-type specific, and this number is even lower after excluding monocytes (12.9%, **Fig. 4a**). Interestingly, these results suggest much higher QTL sharing than those obtained from RASQUAL caQTLs, for which 20,851 (68.5%) were unique to one cell-type, and merely 49 caQTLs were shared in all seven cell-types (**Fig. 4b**). Thus, the discrepancies in sharing between RASQUAL and sc-PME are explained by statistical power, and further increasing QTL mapping power will yield higher estimates of sharing. We conclude that the prevalence of cell-type specific QTLs observed in single-cell genomics compared to bulk genomics data is likely explained by low QTL mapping power rather than cell-type specificity. For this reason, we used sc-PME caQTL results for all analysis below.

**Fig. 4.**
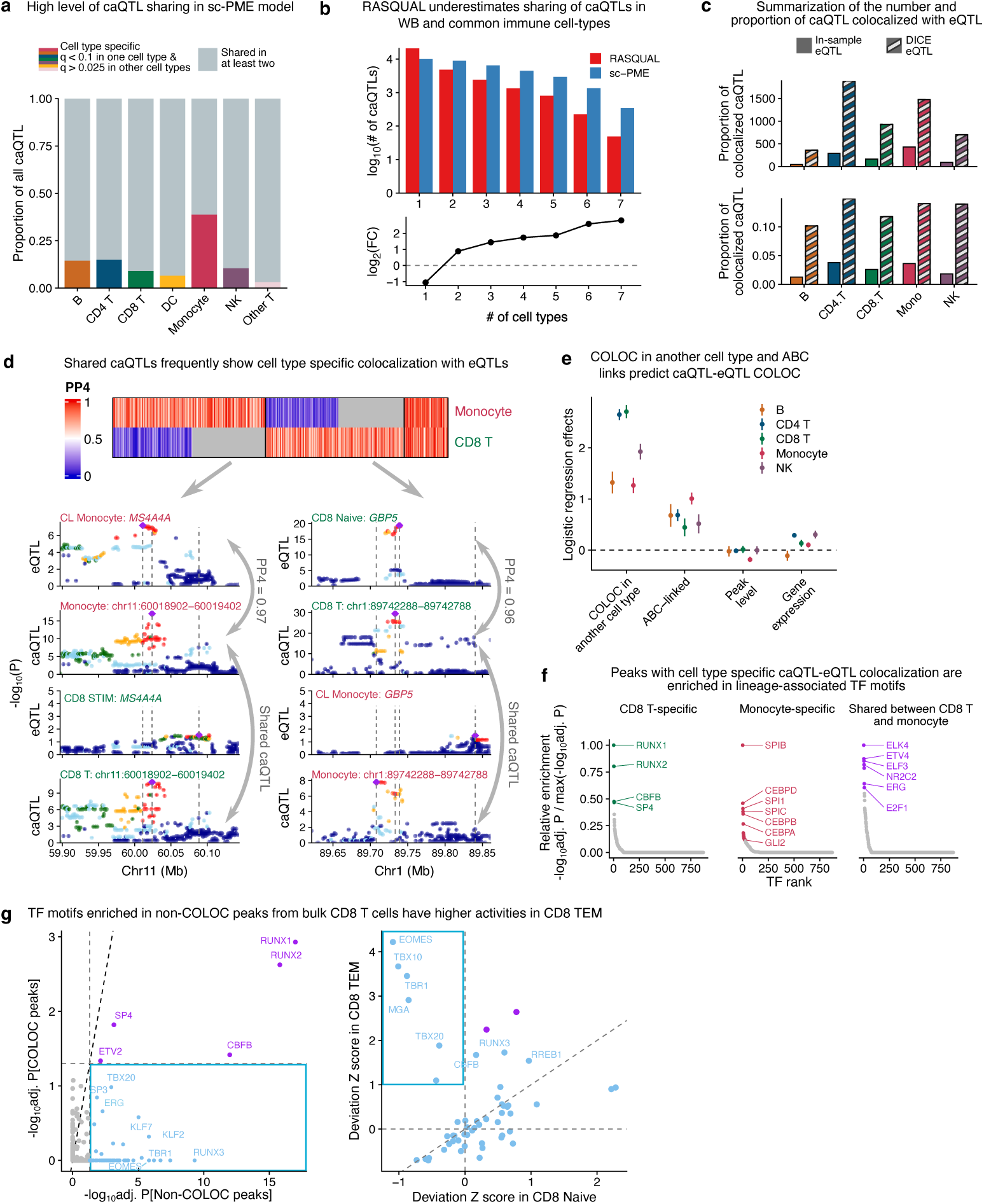
caQTL-eQTL colocalization and dynamic caQTL mapping. **a**, Sharing of caQTLs in sc- PME model in five common immune cell-types. **b**, Comparison of caQTL sharing of caQTLs in seven immune cell-types for RASQUAL and sc-PME model. Top: bar plot of number of caQTLs shared in a given number of contexts. Bottom: log2 fold-change in the number of caQTLs between sc-PME and RASQUAL, highlighting the increased level of sharing for sc-PME caQTLs. **c**, Barplot for the number (top) and proportion (bottom) of caQTLs that colocalize with eQTL from DICE and our in-sample accompanying eQTL data. **d**, Logistic regression coefficients and 95% confidence intervals for variables that predict caQTL-eQTL colocalization. Model includes all caQTL-eQTL pairs that colocalize in at least one cell-type. **e**, Top: heatmap for COLOC PP4 of shared caQTLs between CD8 T cells and monocytes. Bottom: example Manhattan plots for shared caQTL colocalizing with cell-type specific eQTLs in monocytes and CD8 T cells, respectively. **f**, TF motif enrichment in peaks whose caQTL colocalize with eQTL in CD8 T cells and monocytes specifically, or in both cell-types. Colored points represent TFs with adjusted enrichment p-values below 0.05. **g**, TF motifs enriched in non-COLOC peaks in CD8 T cells specifically show higher deviation Z scores in CD8 TEM in our scATAC data.

Chromatin accessibility QTLs are expected to regulate promoter or enhancer activity with an effect on the expression level of one or multiple nearby genes; therefore, we asked whether our sc-PME caQTLs are also eQTLs in the DICE dataset or in the scRNA-seq dataset from matched COVID- 19 patients and controls. As the DICE dataset consist of 15 bulk sorted immune cell types from ∼90 individuals^30^, we matched the 15 cell-types/states available to five cell-types in our scATAC data (excluding DC and “Other T cells” in L1 annotation; **Extended Data** Fig. 5a). In total, we identified 6,228 unique caQTL-eQTL pairs that colocalized in at least one DICE-matching context (referred to as COLOC caQTL-eQTL pairs hereafter), including 2,635 eGenes (24% of all tested) (**Supplementary Table 9**). In comparison, when using our eQTLs from scRNA-seq of matched COVID-19 patients and controls (Randolph et al.), we found fewer caQTL-eQTL colocalizations (representing only 203 cPeaks, or 2.6% of all tested; **Fig. 4c**). This is possibly due to the sparsity of the PBMC scRNA-seq data compared to bulk RNA-seq of sorted immune cells. We therefore used caQTL colocalizations with DICE eQTLs for downstream analyses.

We made two notable observations from this analysis. First, a very small fraction of caQTLs colocalizes with an eQTL. Across five contexts, only 4,088 cPeaks (18.6% of all tested) were colocalized with an eQTL (i.e. “eQTL-caQTL”). This suggests that the majority of caQTLs do not influence the expression level of a nearby gene in the many immune cell-types and contexts included in the DICE dataset (**Fig. 4c**). Second, even when an eQTL colocalizes with a caQTL in DICE, it often does so in just a single context. Indeed, 5,995 (84.5%) of all caQTL-eQTL pairs exist in just one context. This is in stark contrast to the widespread sharing of caQTL we observed. In fact, the majority of caQTLs shared between cell-types do not have shared colocalization with an eQTL. For example, among the 3,091 caQTLs with effects in both CD8 T cells and monocytes, only 134 were eQTL-caQTLs in both cell-types. This number is notably small considering that 4,805 and 7,425 eQTL-caQTL colocalizations were identified in CD8 T cells and monocytes, respectively (**Fig. 4c**). For example, cPeak chr11:60018902-60019402 has a shared caQTL in monocytes and CD8 T cells, but only colocalized with an *MS4A4A* eQTL in monocytes (**Fig. 4d**). Similarly, shared cPeak chr1:89742288-89742788 only colocalized with *GBP5* in naive CD8 T cells in DICE (**Fig. 4d**). In both cases, the lack of colocalization is attributed to the absence of eQTL signals. Thus, we find that eQTL effects are more cell-type specific compared to caQTLs at a large number of loci.

To understand how caQTLs impact gene expression levels in a cell-type specific manner despite affecting accessibility in multiple cell-types, we sought to identify genomic factors that can predict the presence or absence of caQTL-eQTL colocalizations. To this end, we used a logistic regression model to predict caQTL-eQTL colocalization with (1) gene expression level, (2) chromatin accessibility level, (3) enhancer-to-gene links from ABC model^41^ in matched cell-types and (4) whether a caQTL-eQTL pair colocalize in multiple cell-types. Notably, gene expression or chromatin accessibility levels were not predictive of caQTL-eQTL colocalization, suggesting that statistical power does not explain the lack of caQTL effects on gene expression levels. Interestingly, although many caQTL-eQTL colocalizations are cell-type specific, we found that a strong predictor for caQTL-eQTL colocalization in a given cell-type is the presence of a caQTL- eQTL colocalization in another cell-type (**Fig. 4e**), consistent with the notion that *cis*-regulatory elements – particularly promoters – control the same genes across cell-types. We found that the second most predictive feature was enhancer-to-gene links from the ABC model (most predictive in monocyte: log2 effect=1.007, p-value=1.63e-66; least predictive in CD8 T cell: log2 effect=0.44, p-value=4.70e-7), providing a mechanistic explanation for cell-type specific caQTL effects on gene expression levels. These findings highlight the importance of distinguishing “merely active” accessibility peaks from “functional” peaks in caQTL analyses. Critically, our observations indicate that a large fraction of caQTLs have no impact on gene expression levels in any given cell-type because they are not physically connected to any gene in that cell-type.

We also asked whether the presence of specific TF binding motifs could predict eQTL-caQTL colocalization in a cell-type. We focused on a comparison between CD8 T cells and monocytes as many TFs driving accessibility of these two cell-types differ and have been well characterized. Interestingly, we found that, compared to all peaks as background, cPeaks with cell-type specific caQTL-eQTL colocalizations were enriched for known lineage-associated TF motifs, e.g. RUNX1, RUNX2 and CBFB in CD8 T cells, and CEBPA, CEBPB and SPIB in monocytes. By contrast, we found that shared caQTL-eQTL colocalizations are enriched for TFs with broad activity, including ELK4 and E2F1 (**Fig. 4f**). These results suggest that cell-type-specific TFs can potentiate the impact of caQTLs on gene expression in a cell-type-specific manner.

To find potential cellular contexts in which CD8 T cell or monocyte cPeaks without eQTL colocalization might potentiate an eQTL effect, we asked whether cPeaks without any eQTL colocalization were enriched in any recognizable TF binding motifs. In CD8 T cells, we found similar enrichments for a subset of TFs (including RUNX1, RUNX2, CBFB) in cPeaks with or without eQTL colocalizations. However, motifs for another set of TFs (including EOMES, RUNX3 and TBX20) were significantly enriched in cPeaks without eQTL colocalization (**Fig. 4g**, left). Notably, EOMES, RUNX3 and TBX20 are all highly enriched in accessible regions of CD8 TEM cells (**Fig. 4g**, right), supporting a potential gene regulatory function of these caQTLs in CD8 TEM cells. Further supporting this possibility, EOMES regulates differentiation of effector and memory CD8 T cells, and is also implicated in exhaustion; and RUNX factors are associated with CD8 T effector/memory (TEM) populations^42–44^. Although DICE dataset contains eQTLs from 15 different immune cell-types including activated CD8 T cells, it lacks eQTLs from exhausted or long-term memory CD8 T cells. Thus, many cPeaks lacking eQTL colocalizations likely correspond to poised or “primed” enhancers^45^ that can impact gene expression levels in a cell-type-specific manner through activation of cell-type-specific TFs.

Taken together, our findings suggest that the impact of genetic variants on chromatin accessibility can be detected in many cell-types. However, whether chromatin accessibility at these regions influences gene expression levels depends on cell-type specific TFs, resulting in cell-type specific eQTL effects that are difficult to predict from caQTLs alone.

### GWAS loci colocalize more often with caQTL than eQTLs

We sought to characterize the utility of our caQTLs for interpreting immune-related GWAS loci. We colocalized sc-PME caQTLs with GWAS of 11 immune-related diseases, two COVID-19 phenotypes and 36 blood phenotypes. We compared these results with eQTL-GWAS colocalization from DICE^43^. In total, 56.8% (4,696 out of 8,271) of GWAS loci across all traits (GWAS-SNP pairs) colocalized either a caQTL or an eQTL (n=1,532), only caQTLs (n=2,015), or only eQTLs (n=1,149) (**Supplementary Table 10,11**).

Among GWAS loci that colocalizes with both, we found an RA locus^46^ that colocalizes with a *PVRIG* eQTL in Tfh and Tregs, and a caQTL in CD4 T cells. *PVRIG* (also known as *CD112R*) is a co-inhibitory receptor for T cells^47^. The convergence of evidence from gene expression and chromatin accessibility suggests that genetically determined *PVRIG* expression changes underlie RA risks, potentially by modulating T cell activation (**Extended Data** Fig. 5b). As an example of GWAS locus that colocalizes with only eQTLs, *RPS26* gene has a strong eQTL that is shared in all immune cell-types in DICE colocalizing with an RA locus (12:56470625; PP4: 0.93-0.95). Interestingly, the TSS peak of *RPS26* (chr12:56435365-56435865) is accessible in all cell types, but their caQTL did not colocalize with the RA GWAS locus, suggesting possible alternative eQTL mechanisms^48^ (**Extended Data** Fig. 5c).

GWAS loci that colocalize with caQTL but not eQTLs (caQTL-only) are more difficult to interpret. Yet, this category was the largest colocalization category of all, and greatly increased the number of explained GWAS loci from an average of 32.9%, when considering eQTLs alone, to 57.3% when adding caQTLs (**Fig. 5a**). These findings mirror results from recent studies^5,6,49^, which also found that many GWAS loci are associated with changes in chromatin-level phenotypes but not mRNA levels. This led us to wonder how GWAS loci that colocalizes with a caQTL but not an eQTL might function.

**Fig. 5.**
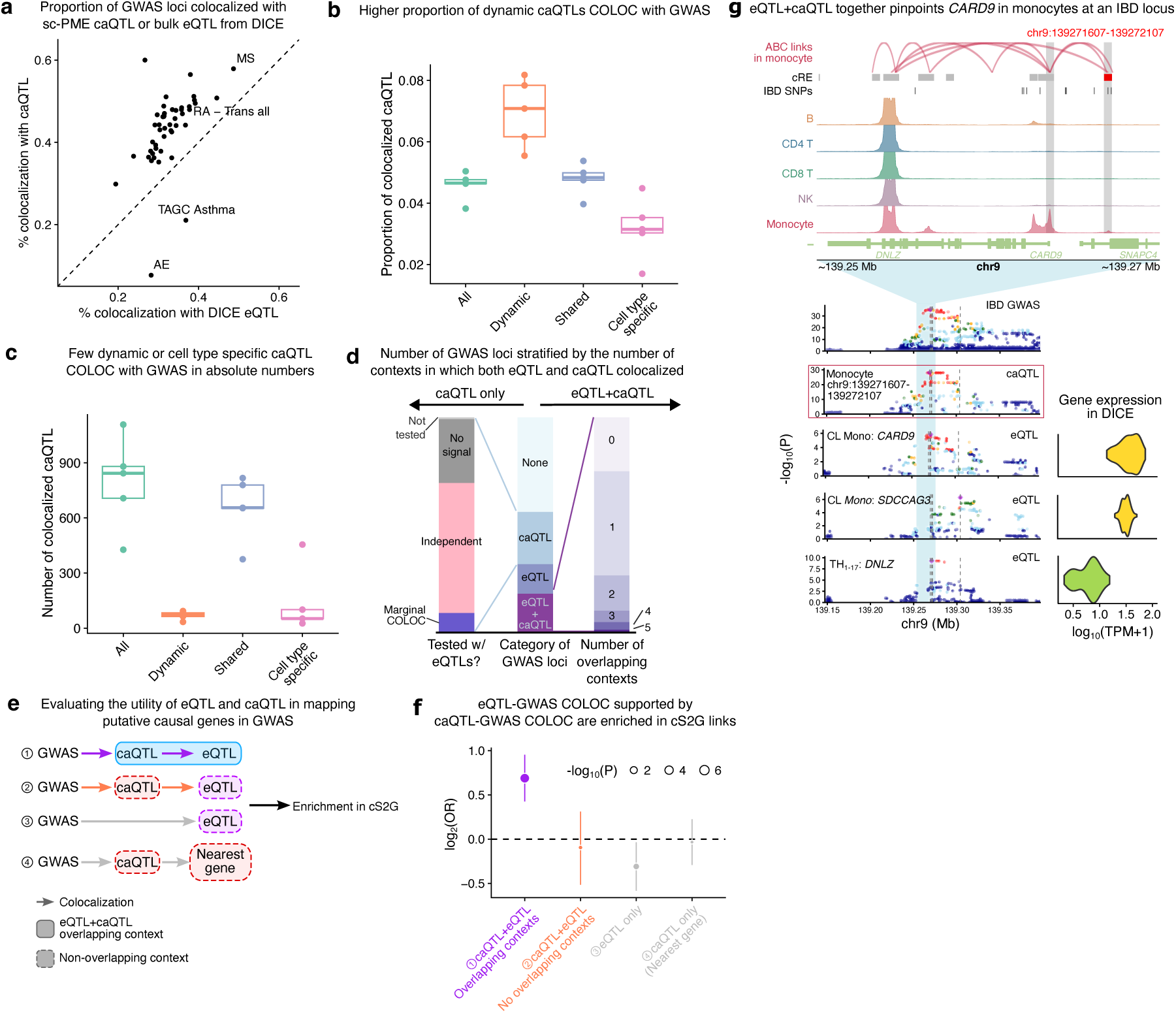
Widespread sharing of caQTLs impedes functional interpretation of disease GWAS. **a**, Scatter plot for the percentage of colocalized GWAS loci with eQTL in DICE and caQTL. **b**, The proportion of caQTL peaks colocalizing with GWAS, categorized by all peaks, dynamic, cell-type specific and shared peaks in each of the five cell-types. **c**, The number of caQTL peaks colocalizing with GWAS, categorized by all peaks, dynamic, cell-type specific and shared peaks in each of the five cell-types. Schematic showing that restricting eQTL-GWAS COLOC by caQTL- GWAS COLOC in the same context to better nominate causal genes and contexts. **d**, Characterization of colocalized GWAS loci by the number of contexts in which they colocalize with either eQTL, caQTL or both. **f**, Forest plot showing that restricting eQTL-GWAS pairs to contexts also supported by caQTL-GWAS COLOC increases enrichment in causal S2G (cS2G) links. Error bars represent 95% confidence intervals of log2(OR) estimates. **g**, An example showing that eQTL- and caQTL-GWAS COLOC together narrows down *CARD9* gene in monocytes as the causal gene and context for an IBD locus. Top: genome browser tracks showing chromatin accessibility around *CARD9* locus. Bottom: Manhattan plots for IBD GWAS, colocalized caQTL and eQTLs in various cell-types. Dashed lines highlight the position of lead SNPs in GWAS and the QTL data. Colored box highlights the nominated causal eQTL.

To nominate causal genes at caQTL-only GWAS loci, we focused on cPeaks overlapping with TSS. In total, 220 (15.3%) GWAS-caQTL loci mapped to promoter regions, which allowed us to nominate a putative causal gene. For example, we identified a peak ∼100 bp upstream of the gene *ZFP36L1*. In CD4 T cells, the same caQTL colocalized with multiple GWAS including CD, ulcerative colitis (UC), RA and MS, suggesting it may affect a common mechanism underlying multiple autoimmune diseases. *ZFP36L1* is an RNA-binding protein that is differentially expressed in osteoarthritis and coeliac disease and has been shown to regulate T cell and B cell development^50–52^, although its exact mechanism in other autoimmune diseases remains unknown (**Supplementary** Fig. 5d). In another case, the promoter peak (chr22:37256806-37257306) for gene *NCF4* has a caQTL colocalizing with an RA GWAS locus in NK and CD8 T cells (**Supplementary** Fig. 5e). *NCF4* has been identified as a risk gene for RA by genetic associations previously; it might be linked to NADPH metabolism in RA and can also regulate NK/CD8 T cell frequencies^53–55^. However, previous eQTL studies did not nominate this gene for RA. These cases suggest that caQTL-GWAS colocalization can, in a limited number of cases, find putative disease genes. The 220 caQTL-only colocalizations gene promoters can be found in **Supplementary Table 12**.

We also asked whether cell-type specific caQTLs or dynamic caQTLs across our defined topic trajectories could explain GWAS hits. On the one hand, we found that cell-type specific caQTLs are less likely to colocalize with GWAS hits compared to shared caQTLs or all caQTLs combined. On the other hand, dynamic caQTLs were more likely to colocalize with GWAS hits than other types of caQTLs, suggesting that caQTLs with dynamic effects tend to capture trait-relevant regulatory elements (**Fig. 5b**). Using the RA GWAS as an example, we found that 24 out 69 (34.8%) GWAS loci that colocalized with a caQTL have an underlying dynamic effect on chromatin accessibility, significantly more than expected by chance (OR=2.27, p-value=1.70e-05; Fisher’s exact test). These results suggest that identifying caQTLs with dynamic effects may help interpret GWAS hits. Still, dynamic caQTLs contributed to very few colocalization with GWAS in absolute numbers because the vast majority of caQTLs do not exhibit dynamic effects, at least at current sample sizes (**Fig. 5c**).

### Limited convergence of caQTL and eQTL signals prevents functional interpretation of most trait-associated loci

We have shown that most GWAS-caQTL colocalizations do not have a corresponding colocalization with an eQTL, and that only a small number of these colocalizations are likely to help with causal gene identification because they impact a gene promoter. We next turned to the GWAS loci that colocalized with both an eQTL and a caQTL and show improved identification of causal disease genes only when both caQTL and eQTL colocalize in the same cellular context.

We first examined the cell-type contexts that were implicated by GWAS loci with both a caQTL and an eQTL colocalization (caQTL+eQTL loci). To allow comparisons between our eQTL and caQTL contexts, we assigned the 15 DICE cell-types to one of the five common immune cell-type contexts defined in our scATAC-seq data (**Extended Data** Fig. 5a). We found that a substantial fraction (375/1,532, 25.4%) of the caQTL+eQTL loci colocalized with the caQTL and eQTL in distinct cell-types (**Fig. 5d**). We also observed that GWAS loci that colocalize with a caQTL or eQTL in multiple contexts tend to colocalize with multiple distinct peaks and eGenes, respectively (**Extended Data** Fig. 6a,b). These observations suggest pleiotropic genetic effects for a substantial fraction of GWAS loci, which hinders our ability to identify which gene or cell context is likely causal for the trait of interest.

We found that 24.4% of caQTL-GWAS colocalizations also colocalize with an eQTL in the same contexts. We hypothesized that eQTLs at these GWAS loci more likely pinpoint the causal disease gene as both caQTL and eQTL converge in the same context. To test this, we leveraged the SNP-to-gene pairs database (S2G)^56^ as a ground truth for causal SNP-to-gene effects. We tested the enrichment of SNP-to-gene links inferred from our QTL data among S2G pairs, using the closest gene as baseline (**Fig. 5e**). We found that caQTL+eQTL S2G pairs in overlapping contexts are significantly enriched for S2G links (log2OR = 0.70, p-value = 7.64e-8; Fisher’s exact test), whereas caQTL+eQTL pairs in different contexts were depleted with S2G links (log2OR=- 0.35, p-value=7.95e-3) (**Fig. 5f**). Furthermore, we found no enrichment in S2G pairs when using the eGene to link SNPs to genes at GWAS loci colocalized only with an eQTL (log2OR=-0.31, p-value=1.89e-2), or when using the nearest TSS to the cPeak at caQTL-only GWAS loci (log2OR=- 0.02, p-value=0.89) (**Fig. 5f**). Thus, GWAS colocalization with a caQTL and an eQTL in the same cell context is helpful for determining causal genes. Conversely and notably, colocalization with a caQTL or eQTL alone or in non-overlapping contexts does not help identify causal genes, at least compared to simply using the nearest gene.

Using GWAS loci that colocalized with a caQTL and eQTL in the same contexts, we were able to narrow down the possible causal genes to no more than two genes and two contexts for hundreds of loci. Many of the candidate genes and cellular contexts have known roles in disease etiology. For instance, one inflammatory bowel disease (IBD) GWAS locus 9:139269198 colocalized with eQTLs of *CARD9* and *SDCCAG3* in monocytes, and that of *DNLZ* in TH1-17 cells. The same IBD locus is also colocalized with two caQTLs in monocytes and NK cells. Thus, we nominated *CARD9* and monocytes as the likely causal gene and context for this GWAS locus. In support of this, *CARD9* (but not *SDCAG3*) is one of the well-established causal genes in IBD^57–59^, and it is the only gene linked to the GWAS SNP 9:139269198 in the S2G database (S2G score=1, the largest possible score in S2G). Furthermore, the colocalized peak is predicted to be connected to *CARD9* TSS in monocytes in the ABC model, whereas no enhancer-TSS links were found in T cells. *CARD9* is highly expressed in classical and non-classical monocytes, whereas *DNLZ* is lowly expressed in TH1-17 cells. The identified peak (chr9:139271584-139272084, ∼13.3Kb upstream of *CARD9* TSS) is also monocyte-specific and harbors fine-mapped IBD GWAS SNPs (**Fig. 5g**). This example highlights the need to consider multiple lines of molecular information to obtain a set of high-confidence targets, and supports our interpretation that caQTL or eQTL colocalization can be misleading.

In conclusion, GWAS colocalization with both a caQTL and an eQTL gives us the best chance at identifying the causal genes and contexts underlying trait association. However, most caQTL- GWAS colocalization identified in our study did not colocalize with an eQTL in any context. Thus, although genetic effects on chromatin accessibility across multiple immune cell-types and contexts drastically increase GWAS loci colocalization rates to a molecular QTL, our work suggest that they should be interpreted with caution as their effects on gene expression levels are yet unidentified.

## Discussion

In this study, we constructed a harmonized map of single-cell chromatin accessibility accompanied with high-quality genotype information. Unlike single cell eQTLs studies which have become streamlined with the development of new approaches and best practice^60,61^, there exists few studies and approaches for caQTLs mapping in single cells. We show that by applying both allelic-imbalance modeling in RASQUAL and the sc-PME model, we can identify a set of high-confidence caQTLs whose summary statistics can be used for state-of-the-art downstream analyses. In total, we were able to identify 37,390 caQTLs from our integrated scATAC-seq dataset, which quadrupled the number of caQTLs from that reported in a recent, smaller study^36^. We also utilized the continuous nature of cell loadings in topic analysis to map dynamic caQTLs. We find that topics are more straightforward to interpret compared to principal components^36^.

Remarkably, we found that adding caQTLs increases colocalized GWAS loci by an average of ∼50% compared to eQTLs alone, suggesting that mapping caQTLs in single cells may be a promising paradigm for studying and biologically interpreting disease associated variants. As such, our work corroborates similar findings reported from recent bulk caQTL studies^42^.

Still, there has been no straightforward explanation as to why, in any given cell-type context, many GWAS loci only have effects on chromatin, but not on gene expression level. We found that the sharing of caQTLs across immune cell-types is widespread, but their impact on gene expression levels is much more restricted owing to cell-type specific gene expression and/or cell-type specific enhancer-promoter interaction. Thus, we interpret novel caQTL-GWAS colocalization results with caution. We posit that many – if not most – caQTL-GWAS colocalizations do not reflect meaningful regulatory effects in a causal cell-type context. Thus, finding the gene and cell-type context that causally mediates genetic effects on complex traits may be difficult even when a caQTL-GWAS colocalization has been found. Our findings that caQTLs are widely shared across immune cell-types and states indicate that most of these caQTLs may impact gene expression levels in immune contexts for which eQTLs are unavailable. Thus, increasing the cell context coverage maps of eQTLs will help in finding more cell-type contexts in which caQTL, eQTL, and GWAS signals all align, but this will also lead to uncertainty in which context is most relevant for the trait.

Our findings are consistent with the existence of poised or primed enhancers, which are accessible in many cell-types and states, but affect expression level of nearby genes only when context specific transcription factors are activated (e.g. upon immune stimuli or during differentiation)^45^. However, the concept of primed enhancer does not generalize well across very different immune cell-types such as B cells and monocytes, in which many open chromatin regions are shared, but their impact on gene regulation may differ. Of note, a recent multi-modal study shows that open chromatin states are shared among various RA-associated cell-states, whereas the gene expression profiles are distinct^62^. More work is required to figure out how the same *cis*-regulatory region can impact different genes in different cell-types. But what is clear is that these effects appear prevalent and complicates fine-mapping of causal genes at disease GWAS loci.

In conclusion, we demonstrate the utility of chromatin accessibility data in the functional study of regulatory elements, but we argue that caQTL data — especially those without clear effects on gene expression levels — often do not have any downstream regulatory impact. Thus, we highlight the need for integrating additional and orthogonal information such as eQTL data, enhancer-promoter links (e.g. ABC score), and other functional data to elucidate mechanisms of GWAS loci. Our findings suggest that population-scale studies using multi-omic single cell assays in disease-relevant contexts will be instrumental in improving our ability to uncover the molecular mechanisms through which individual non-coding variants impact disease risk. More challengingly, our results suggest that we have yet to identify many of the causal cell-types for GWAS traits. Mapping eQTLs in relevant cell populations from disease patients^1,63,64^ or from organoids^65,66^ subject to diverse treatments may be a fruitful approach in this direction.

## Methods

### Clinical sample collection

We collected PBMCs from 20 COVID-19 patients hospitalized in Montréal, Canada with COVID- 19 between April 2020 and December 2021 who initially presented with symptomatic, primary infection. All acute phase samples were collected from unvaccinated patients within 20 days of symptom onset. None received plasma transfer therapy. We also prospectively sample a subset of the same patients during convalescent phase COVID-19 (n = 11), and healthy controls (n = 5). The respective institutional IRBs approved multicentric protocol: MP-02-2020-8929. Written, informed consent was obtained from all participants or, when incapacitated, their legal guardian before enrollment and sample collection.

### scATAC-seq sample processing

#### Initial preparation

Cryopreserved samples were thawed and cultured in RPMI 1640 without glutamine (Fisher) supplemented with 10% fetal bovine serum (Corning), 1% L-glutamine (Fisher), and 0.01% gentamicin from 10 mg/mL stock (Fisher) overnight. After incubation, samples were washed with PBS, passed through a 40μm filter, and manually counted with trypan blue staining by brightfield hemocytometer.

#### Nuclei isolation

As in the demonstrated protocol CG000169, Rev. E, from 10X genomics, we lysed each batch of 1 million cells with a IGEPAL 630 / digitonin based lysis buffer for 3 minutes on ice. Nuclei were then washed once and resuspended in 7.5uL of 10X Genomics Nuclei Buffer. Finally, 2.5uL of nuclei were counted by trypan blue staining on a hemocytometer.

### Single-cell ATAC library preparation and sequencing

#### Single-cell ATAC capture

We followed the 10X Genomics Chromium Next GEM Single Cell ATAC Reagent kit with version #1.1. Nuclei were transposed by isothermal incubation at 37°C and Post Gel Bead-in-Emulsion (GEMs) were generated by the 10X Controller and subjected to PCR as described in the 10X User Guide, and post-incubation products were stored at -20°C until downstream processing. Each sample was captured individually (i.e. without pooling) aiming for 5,000 nuclei from each sample to be captured.

#### Single-cell ATAC library preparation

Post GEM incubation cleanup and sequencing library preparation were performed as described in the Single Cell ATAC Reagent Kits v1.1 User Guide (10X Genomics). Briefly, we cleaned up post-incubation GEMs first with DynaBeads MyOne SILANE beads (ThermoFisher Scientific) and then with SPRIselect reagent (Beckman Coulter). Libraries were constructed by performing sample index PCR (98 °C for 45 s, 9 or 10 cycles of 98°C for 20 s, 67°C for 30 s, 72°C for 20 s, and 72°C 1 min) followed by SPRIselect size selection.

#### Next-generation sequencing

Prior to sequencing, all multiplexed single-cell libraries were quantified using the KAPA Library Quantification Kit for Illumina Platforms (Roche) and pooled in an equimolar ratio. Libraries were sequenced by 100 base pairs (read1: 50, i7: 8, i5: 16, read2: 50) on an Illumina NovaSeq 6000.

### Preprocessing of in-house and public scATAC-seq data

We processed data in all three studies from FASTQ files using the following pipeline. Reads were processed using cellranger-atac v2.1.0 with an in-house GRCh37 reference genome generated using scripts from 10X Genomic documentations (https://support.10xgenomics.com/single-cell-atac/software/release-notes/references#GRCh38-2020-A-2.0.0). We removed reads that were unmapped, did not have primary alignment, failed platform/vendor quality checks, and had duplicated or supplementary alignment; we only kept reads that were paired and mapped in proper pairs (‘samtools view -f 3 -F 3844’). We then removed allelic-biased reads using the WASP^67^ workflow implemented in Hornet. We converted the resulting BAM file from each library into a fragment file using sinto v0.7.5 and loaded into an ArchR project separately. We then analyzed each library separately to identify high-quality barcodes and remove doublets. As a first pass, we excluded cell barcodes with fewer than 1,000 and more than 50,000 unique fragments, with a TSS enrichment score lower than six for all libraries and excluded those with high ratios of reads mapping to nucleosomes, mitochondrial genome or ENCODE blacklist regions in a library-specific manner (**Extended Data** Fig. 1b). We also used AMULET^68^ on BAM files to flag and remove potential doublets (AMULET q-value < 0.1).

### Preprocessing and cell-type annotation of public scRNA-seq data

We re-analyzed previously published PBMC scRNA-seq data from COVID-19 patients^67^. Count matrix was downloaded from Human Cell Atlas webpage and converted to a Seurat object. As a first pass, we ran Azimuth with PBMC reference to annotate all the cells. We compared Azimuth L1 annotation (B, CD4 T, CD8 T, dendritic cells (DC), monocytes, natural killer cells (NK), other T cells) with original cell type labels provided by the author, and only kept cells with consistent labels.

### Basic analysis of scATAC-seq data

Through the processing steps above, we identified a list of barcodes that represent high-quality single cells with individual ID for each library. We then loaded the fragment files containing these barcodes from all libraries to one ArchR project for integrated analysis. Dimension reduction on this full dataset was performed on the binary tile matrix, selecting the top 30,000 variable tiles and outputting 50 reduced dimensions with ’addIterativeLSI’ function in ArchR. We then feed this LSI projection to the ’reducedMNN’ function in R package ’batchelor’ to remove batch effects across libraries^69^. We implemented a wrapper function to add MNN-adjusted dimensions to the ArchR project object, enabling downstream analysis within ArchR framework. Cell clusters were identified with a resolution of 0.8. For visualization, the reducedMNN-adjusted LSI was used to derive a UMAP embedding with ‘minDist=0.8’ and ‘spread=1’.

To calculate gene activity scores (GA scores) from scATAC-seq profiles, we generated an in-house gene reference set from the GENCODE v19 annotation. Basically, we started from all the gene symbols in the full GENCODE annotation and removed those whose ‘gene_type’ map to one of the following: snRNA, misc_RNA, snoRNA, rRNA, miRNA, pseudogene, polymorphic_pseudogene, IG_V_pseudogene, TR_V_pseudogene, IG_C_pseudogene, TR_J_pseudogene, IG_J_pseudogene, processed_transcript, sense_intronic, 3prime_overlapping_ncrna and sense_overlapping, keeping 32,885 genes on chr1-22 and chrX. We then extracted the transcript start sites (TSS) and exons for these genes and constructed a gene annotation object that was added into our ArchR project. Our custom annotation includes important marker genes that are missed in the default hg19 annotation used by ArchR, such as gene *LINC02446* (also known as *RP11-291B21.2*, **Fig. 1f**), a long non-coding RNA that marks activated CD8 T cells^9^. Using this custom gene annotation, we then calculated the GA score using ‘addGeneScoreMatrix’ with default parameters in ArchR.

To better annotate cell-types in our scATAC-seq data, we integrated it with our Azimuth-annotated scRNA-seq data and transferred the annotation labels to scATAC-seq cells. We first performed unconstrained integration using the ‘addGeneIntegrationMatrix’ function in ArchR. We then examined the confusion matrix between cell clusters and annotated cell-types. Several clusters contained mixed cell-types from the reference dataset. Upon further speculation, we found these clusters tend to have higher rates of mitochondrial DNA and lie between well-defined cell-types in the UMAP, suggesting these cells are of lower quality or are potential unremoved doublets. We excluded these cells from the dataset and performed constrained integration by restricting cells within four groups: T/NK cells, monocytes/DC, B cells and other (platelet and HSPC). After this round of constrained integration, we found several T/NK cell subtypes in L2 have very low cell numbers in scATAC-seq data, we therefore only kept labels with sufficient cell numbers (CD14 Mono, CD16 Mono, NK, NK_CD56bright, NK Proliferating, pDC, cDC2, Platelet, B naïve, B intermediate, B memory, Plasmablast, gdT, MAIT, CD4 Naïve, CD4 TCM, Treg, CD8 Naïve, CD8 TEM, CD8 TCM, HSPC) and performed another iteration of constrained integration. Finally, we re-calculataed the LSI, MNN-adjusted dimensions and the UMAP embedding and Leiden clustering on the remaining cells.

To identify candidate peaks, we first produced pseudo-bulk group coverages in each Leiden cluster and used the three studies as sample labels in ‘addGroupCoverages(sampleLabels=“Sample”)’. We then called reproducible peak set by setting ‘reproducibility=2’ in ‘addReproduciblePeakSet’. In this way, we were able to identify peaks that are called in at least two of the three studies in our data.

### Genotype imputation from aggregated scATAC-seq data

Read coverage across the genome was visualized using ’plotCoverage’ from deepTools^70^, excluding blacklist regions from ENCODE. Genotype likelihood calculation and imputation were performed following GLIMPSE documentation^69^. Briefly, we first inferred genotype likelihoods across all SNPs in the 1000 Genome Project from filtered BAM files from scATAC data with ’bcftools mpileup’. In this step, we only included sites with sequencing depth below 15 (“bcftools view -i ’FORMAT/DP<=15’”) to avoid regions with unreasonably high read coverage. Next, we merged genotype likelihood from all individuals from the three studies and performed GLIMPSE genotype imputation jointly. Imputed genotypes were phased with eagle v2.4.1^71^.

To confirm GLIMPSE-imputed genotypes from scATAC-seq reads are of high quality and are not biased by strong allele-specific signals in accessible chromatin regions, we compared Minimac4 imputation from microarray with GLIMPSE results in the 13 individuals with microarray data from Benaglio et al^7^. We first imputed the microarray genotype data from the original study using the pipeline documented in Michigan Imputation Server^72^ (https://imputationserver.readthedocs.io/en/latest/pipeline). We used the same reference panel as in our GLIMPSE pipeline. We then calculated mean imputation quality score (INFO score) for SNPs stratified by reference MAF bins. We also calculated correlation between genotype dosages for SNPs imputed using GLIMPSE and Minimac4 and derived mean correlation across the 13 individuals in each reference MAF bin using ‘vcf-stats’.

### Topic modeling on scATAC count data

#### Fitting the topic model

Topic modeling was performed using the R package ‘fastTopics’^71^. We retrieved the cell-by-peak count matrix from the ArchR object. In practice, we considered two aspects in fitting Poisson NMF to our count data. First, fitting the topic model on the full data is computationally expensive. Second, the NMF problem is non-convex, meaning that each model fit returns slightly different results, making it difficult to compare the output using different parameters even on the same data. To speed up the model fitting process, we randomly down sampled 10,000 cells. For peaks that have zero counts in these 10,000 sampled cells, instead of removing them from the matrix, we further sampled cells where they have non-zero counts. This ensures that we can project the fitted model to the full count matrix. In total, 10,711 cells were used for the initial model fitting. To make sure we can easily compare multiple model fits on the same data, we first performed NMF using a small number of total topics (k), and then fit NMF with more topics conditioning on the previous model fit. We started by fitting the topic model with k=6 using the ‘fit_topic_model’ function, using 100 main iterations and 200 refining iterations (‘numiter.main=100, numiter.refine=200’). This returned a multinomial topic model fitting, which was then projected to the full count data using the ‘predict’ function implemented in ‘fastTopics’. To fit a model with eight topics (k=8), we propagated the loading matrix and the factor matrix from k=6 with two more columns of uniformly distributed values (1/k for loading matrix and 1/[number of peaks] for score matrix). We then applied the fitting steps adapted from the ‘fit_topic_model’ function. Breifly, the expanded loading matrix and factor matrix were passed into ‘init_poisson_nmf’ together with the down sampled count matrix to initialize a new Poisson model. Then, the model was fitted with EM algorithm for 100 iterations (main fitting) and updated with SCD algorithm for 200 iterations in two consecutive runs of the ‘fit_poisson_nmf’ function. The fitted Poisson model was converted to a multinomial NMF model with ‘poisson2multinom’ function. The output of this final step is a cell-by-eight-topic loading matrix. We iterated this process with 10, 12, 14, 16, 18 and 20 topics. This framework allowed us to keep the order of topics constant, making it possible to compare across model fits, while updating them when more topics are added. To visualize topic modeling results in a Structure plot, we performed PCA on the loading matrix (after centering and scaling) and used the rotated data matrix for K-means clustering (K=30). To avoid over-plotting, we randomly selected 2.5% of cells from each cluster, resulting in a subset of ∼5,500 cells for visualization.

#### Calculation of gene-level scores from peak-level scores

To define a molecular program underlying each topic, we relied on the factor matrix. We selected the top 10% of peaks with the highest score in each topic. To calculate gene-level scores from peak-level scores, we applied ArchR’s exponential-weighting strategy to calculate gene activity scores. Briefly, scores of peaks within the gene body are directly summed up, and scores of peaks up to 5 Kb upstream of the gene TSS are weighted by distance-based power-law. We then calculated the Z score of each gene across all the topics.

#### Stratified-LDSC analysis on top peaks in each topic

For s-LDSC analysis, we selected top 10% peaks with the highest scores from each topic. Each peak was extended to 1,500 bp around its center. We used GWAS summary statistics for 50 phenotypes (11 immune-related diseases and 36 blood cell-type traits, and height as a negative control) (**Supplementary Table 13**). GWAS summary statistics were munged by ‘munge_sumstats.py’. LD score calculation and s-LDSC analysis were carried out according to LDSC documentation conditioning on default baseline annotations.

#### Association between topic k17 loadings and COVID-19 status

To test whether k17 loadings are associated with COVID-19 status, we first calculated average donor-level k17 loadings for each sample. We only included cells from healthy controls or active COVID-19 at the time of sample collection and with k17 loading larger than 0.01, as cells below this cutoff largely represent estimation noise. We then fitted two mixed-effects logistic regression models:

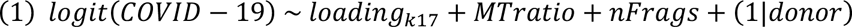

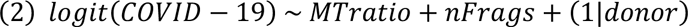

Formula (1) is the full model and (2) is the null model. We then performed a likelihood ratio test (LRT) with ‘anova’ function in R to test whether k17 loadings significantly predicts donor COVID- 19 status.

#### Trajectory analysis in topic model

We defined the cell trajectory directly from topic loadings with slight modifications to accommodate the analysis workflow of the ArchR package. As a proof-of-concept, we first scrutinized the B cell trajectory. Since k2 represents naive B cells, its loadings are the highest in naive B cells and decreases in memory B cells and plasmablast. To construct a trajectory that represents B cell maturation, we used the reverse order of k2 loadings, such that the trajectory value increases as naive B cells transit into memory B cells. The trajectory was restricted to L1-annotated B cells, and we set the value of all other cell-types to ‘NA’, so that ArchR does not use these cells in the analysis. The trajectory values were then scaled to the range of 0-100 for downstream analysis. To study the change in the proportion of memory B cells or plasmablasts along the trajectory, the trajectory was divided into percentiles, and we calculated the proportion of non-naive B cells in each percentile according to L1 annotation. When building a trajectory for k9, we removed cells whose k9 loadings are below 0.1, as these likely represent background noise rather than biologically meaningful variations.

After deriving the trajectory values from cell loadings, we added the trajectories to the ArchR project as a metadata column. To visualize the changes of gene activity scores along the trajectory, we used the ‘getTrajectory’ function, followed by ‘plotTrajectoryHeatmap’ functions with options ‘varCutOff=0.8, returnMatrix=TRUE’. The heatmap was visualized using ComplexHeatmap^73^.

To assess the relevance of k17 trajectory to COVID-19, we first asked in a cluster-based analysis, how many differentially active genes can be found between healthy and COVID-19 cells. To do so, we compared all COVID-19cells with all healthy cells in k17 trajectory regardless of k17 loadings using the ‘getMarkerFeatures’ function. We then grouped cells into quintiles according to their k17 loadings, where higher quintiles were enriched for more COVID-19 cells. We next tested for differential gene activity between all cells in the first quintiles and COVID-19 cells in the higher quintiles (second and above). Note in this test we used healthy and COVID-19 cells in the first quintile as control, following the idea that COVID-19 cells in the first quintile are epigenetically similar to healthy cells.

### Chromatin accessibility QTL mapping

#### RASQUAL caQTL mapping

Chromatin accessibility QTL (caQTL) were first mapped using RASQUAL on three grouping levels, whole blood-like (WB-like), L1 and L2 annotations. We generated pseudobulk counts by summing single-cell counts across cell barcodes within each group. For the WB-like group, we included all peaks and all individuals. For caQTL mapping in L1 and L2 cell-types, we only included cell-types with at least 50 cells in at least 10 individuals. From the pseudobulk count table, we calculated library sizes and phenotype PCs (after scaling and centering; using ’prcomp’ function in R). To get allelic-specific read counts, we extracted reads from each group using ’filterbarcodes’ command from sinto v0.7.5 and counted allelic-specific reads using ’createASVCF.sh’ from RASQUAL. We only kept bi-allelic SNPs with at least four minor allele counts across tested individuals. We included library size as offsets and five genotype PCs, the number of cells, and the GC content for each peak as covariates. RASQUAL was run in nominal mode and permutation mode. We extracted the lead SNP for each tested peak and used nominal and permuted log10(q-value) from RASQUAL to calculate empirical p-values with ‘empPvals’ function from the ‘qvalue’ R package, and then derived q-values from the empirical p-values. We used q-value below 0.1 as the cutoff for significant caQTLs.

#### Enrichment of RASQUAL caQTLs in bulk LCL caQTLs

To test the enrichment of RASQUAL caQTLs from WB in bulk caQTLs from LCL, we obtained summary statistics from a previous study^28^. We extracted lead SNP for each peak in the LCL dataset and ranked them by their significance. We then tested the enrichment of RASQUAL lead SNPs in the top 1%, 5%, and 10% of the most significant LCL caQTL lead SNPs using Fisher’s exact test.

#### Enrichment of RASQUAL caQTLs in DICE bulk eQTLs

To test the enrichment of RASQUAL caQTLs from common immune cell types in bulk eQTLs from DICE, we first extracted the genomic locations of lead eQTL SNPs and extended it by 500 bp on each side and converted it to bed format. These eQTL regions form the genomic annotation in which caQTL enrichment is tested. We then converted RASQUAL lead caQTL SNP positions to bed format, and tested their enrichment in eQTL regions using QTLtools ‘fenrich’ command^74^, while feeding our peak set into the ‘--tss’ argument to adjust for the fact that RASQUAL caQTLs are enriched in the cPeaks themselves. The odd ratios from ‘fenrich’ were visualized after being normalized to the maximum odd ratio in each caQTL cell type.

#### Single-cell Poisson mixed-effect model (sc-PME) caQTL mapping

Single-cell caQTL mapping with the sc-PME model was first performed in three studies separately. For continuous covariates, we included top five genotype PCs, top five LSI dimensions, TSS enrichment scores, fraction of mitochondrial reads, log10 of number of unique fragments, all of which are scaled and fitted as fixed effects. We also included libraries and donors as random effects. The Poisson mixed effect model was fitted using the ‘glmer’ function (‘family=poisson’) in the lme4 R package. We set the additional options as ‘nAGQ=0, control=glmerControl(optimizer=“bobyqa”, calc.derivs=F)’ to save computational time for model fitting. We performed meta-analysis using effect sizes and standard errors from all SNPs in the three datasets and ran Metasoft without genomic control. For downstream analysis, we used effect sizes and standard errors from the random effects model and p-values from the Han and Eskin’s Random Effects model (RE2)^34^. To call significant caQTL in the meta-analyzed sc-PME results, we first applied Bonferroni correction for all SNPs in a given peak, extracted the lead SNP for each peak, and then calculated q-values from the Bonferroni-adjusted p-values across all lead SNPs. We used a q-value below 0.1 as a cutoff for significant lead caQTL.

#### Dynamic caQTL mapping with sc-PME

To identify dynamic effects of lead caQTL SNPs along cell trajectories defined by topic modeling, we tested for interaction between genotype dosages and topic loadings. To avoid confounding dynamic caQTL with cell-type specific caQTL, we (1) mapped dynamic caQTLs separately in each common cell-type and (2) only included topics that are present in each cell-type as follows: B (k1, k11), CD4 T (k6, k7, k17), CD8 T (k3, k6, k7, k14, k17, k18, k19), NK (k3, k17), monocyte (k10, k12, k15, DC (k4, k10, k12, k15), other T cell (k3, k6, k8, k14, k17, k18, k19). For each SNP, we fitted a model with genotype-by-loading interaction term and a reduced model without the interaction term. We then use R function ‘anova’ to perform a likelihood-ratio test (LRT) comparing the two models, and used p-values from LRT to call significant dynamic caQTLs. Because we only tested the top caQTL SNP for every cPeak, we calculated q-values from LRT p-values in each topic separately, and then multiplexed the q-value by the number of topics in which a given SNP was tested for; this is equivalent to a Bonferroni adjustment on the number of topics tested for each SNP, thus caQTL SNPs from cells with more topics (e.g. CD8 T cells) were subjected to more stringent significant level cutoff. We reported adjusted q-value below 0.01 as significant dynamic caQTLs.

### Colocalization of caQTLs with bulk eQTLs and GWAS

To perform colocalization between caQTLs and eQTLs, we used the eQTL summary statistics from the DICE study we re-processed and published before^1^. We tested for colocalization between a caQTL and an eQTL when there are more than 150 overlapping SNPs and their corresponding lead SNPs are among the overlapping SNPs.

To perform colocalization between our caQTL and in-sample COVID-19 eQTLs, we used the list of significant eGenes defined as mashr^34^ local false sign rate (lfsr) below 0.1 in the accompanying manuscript (Randolph et al.). We tested for colocalization between a caQTL and an eQTL when there are more than 150 overlapping SNPs and their corresponding lead SNPs are among the overlapping SNPs.

To perform colocalization between caQTL and GWAS, we used GWAS summary statistics of 11 immune-related diseases and 36 blood cell-type GWAS that we accessed previously^1^. Briefly, we defined a GWAS locus as a 1 Mb window centered around a SNP with a p-value below 1e-7, starting from the SNP with the smallest p-value, removing all SNP lies within 1 Mb, and iteratively identified all GWAS loci until no SNPs with p-value below 1e-7 remained. Like eQTL, we tested for colocalization between a caQTL and a GWAS locus when their corresponding lead SNPs are among the overlapping SNPs and there are more than 150 overlapping SNPs. All colocalization analyses were performed with the ‘coloc.abf’ function from R package ‘coloc’ v5.2.1^30^.

## Data Availability

All data produced in the present study are available upon reasonable request to the authors after final publication of the article.

## Acknowledgements

We thank N. Gonzales for her careful reading of our manuscript and their insightful comments. This work was completed in part with resources provided by the University of Chicago Research Computing Center. This work was supported by National Institute of Health grants R35GM153249 (Y.I.L.), R01GM130738 (Z.M, Y.I.L), R01GM134376 (L.B.B.) and R35GM152227 (L.B.B.). This work was also supported by Canadian Institutes of Health Research grants VR2-173203 and 178344 to D.E.K. We also acknowledge the support from the UChicago DDRCC, Center for Interdisciplinary Study of Inflammatory Intestinal Disorders(C-IID) (NIDDK P30 DK042086). H.E.R. was supported by a Ruth L. Kirschstein National Research Service Award (NHLBI F31-HL156419). The Biobanque Québécoise de la COVID-19 (BQC19) is supported by the FRQS Génome Québec and the Public Health Agency of Canada. L.B.B. and Y.I.L. are CZ Biohub Investigators.

## Author contributions

Y.I.L. and L.B.B. jointly supervised research. Z.M., Y.I.L, and L.B.B. conceived and designed the experiments. H.E.R, E.K., A.D., V.L., C.B, D.E.K performed the experiments. Z.M. performed statistical analyses and analyzed the data. H.E.R. and R.A.-G. also performed data analysis. Z.M. and Y.I.L. wrote the paper with critical contributions from L.B.B. and X.L. and input from all authors.

---

**Extended Data Fig. 1.**
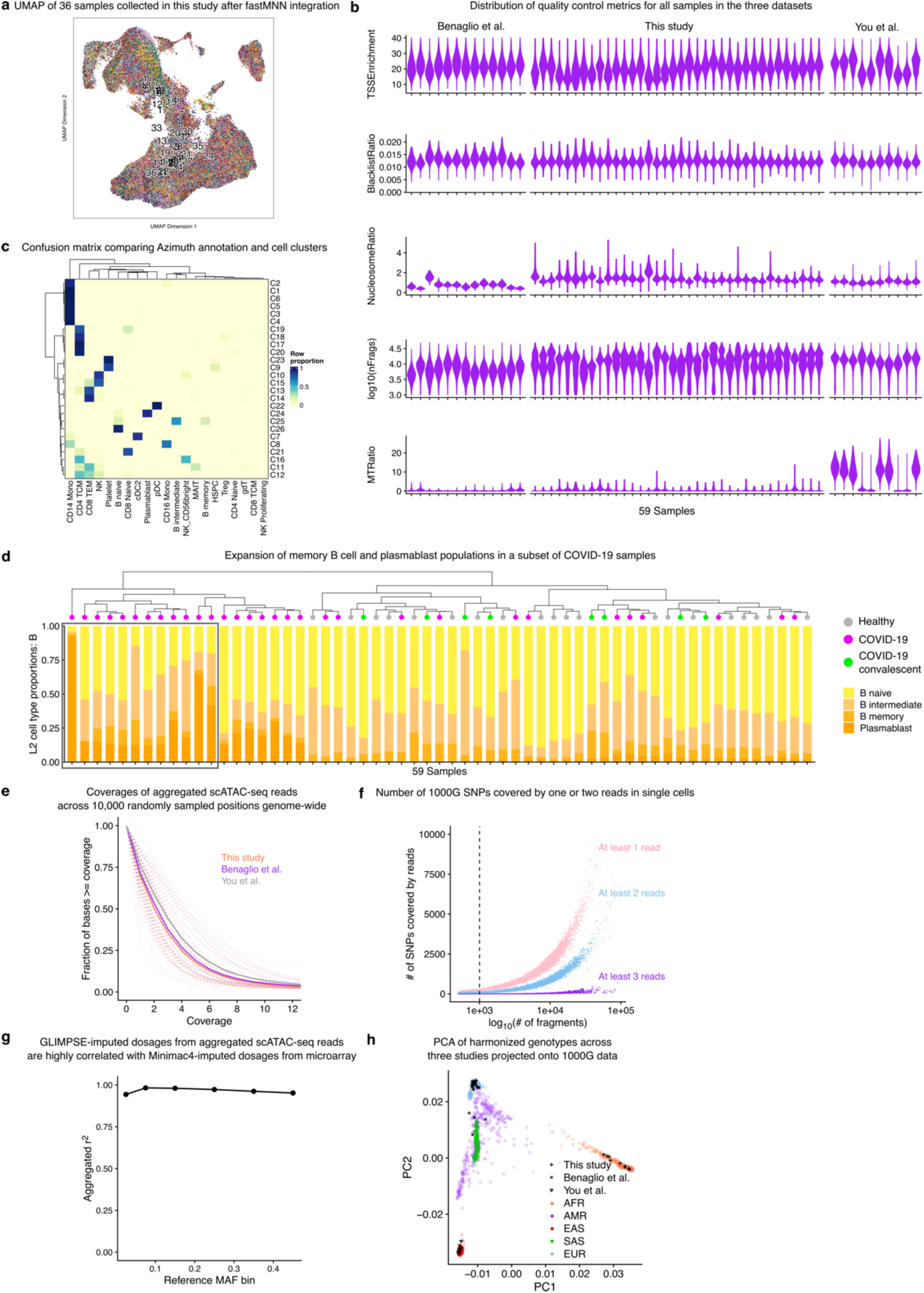
Quality control on scATAC-seq data and genotyping. **a**, UMAP of integrated 36 samples collected in this study. **b**, Violin plots showing the distribution of common quality control metrics for all cells in the three datasets. **c**, Confusion matrix comparing Azimuth L2 annotation and cell clusters. Heatmap is colored by row normalized proportions, such that the cell-type compositions sum up to one on each row. **d**, Bar plots showing expansion of memory B cells and plasmablasts (highlighted in border) in a group of COVID-19 samples. **e**, The three scATAC-seq data sets used in this study have very similar read coverages at 10,000 randomly sampled positions genome-wide. **f**, Number of 1000G SNPs covered by at least one, two or three reads in each single cell as a function of the number of unique fragments. Dashed line indicates 1,000 unique fragments, the cutoff we used for filtering low-quality cells. **g**, Mean correlation between GLIMPSE-imputed genotype dosages from aggregated scATAC-seq reads and those imputed from microarray data using Minimac4. **h**, PCA analysis of all individuals in this study with 1000G samples.

**Extended Data Fig. 2.**
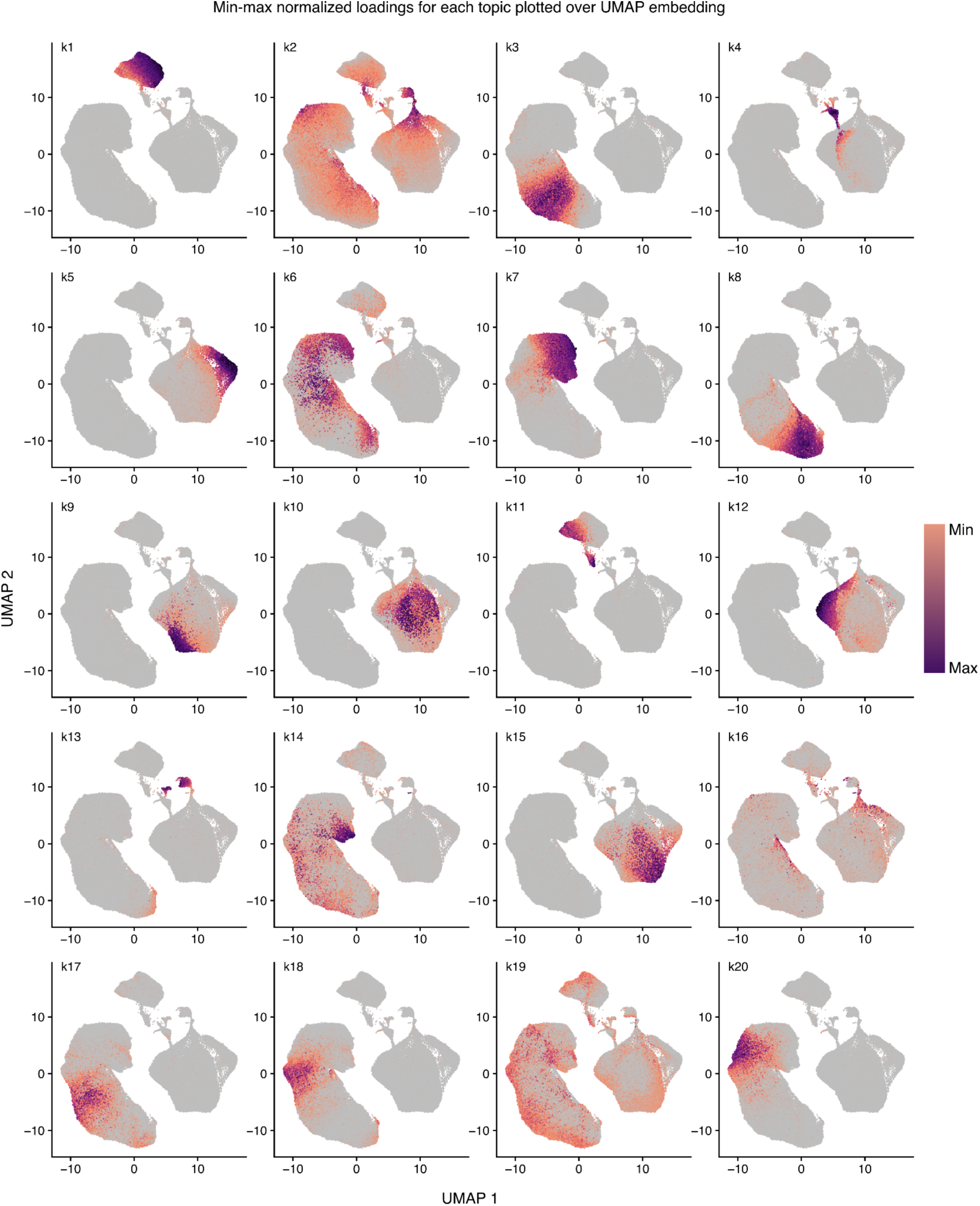
Visualization of cell loadings for the 20 topics in UMAP embedding. UMAP plots showing the distribution and quantity of loadings for the 20 topics. Loading scores are normalized by min-max for each topic, and loading scores below 0.05 in a cell were set to 0 for visualization purposes (gray).

**Extended Data Fig. 3.**
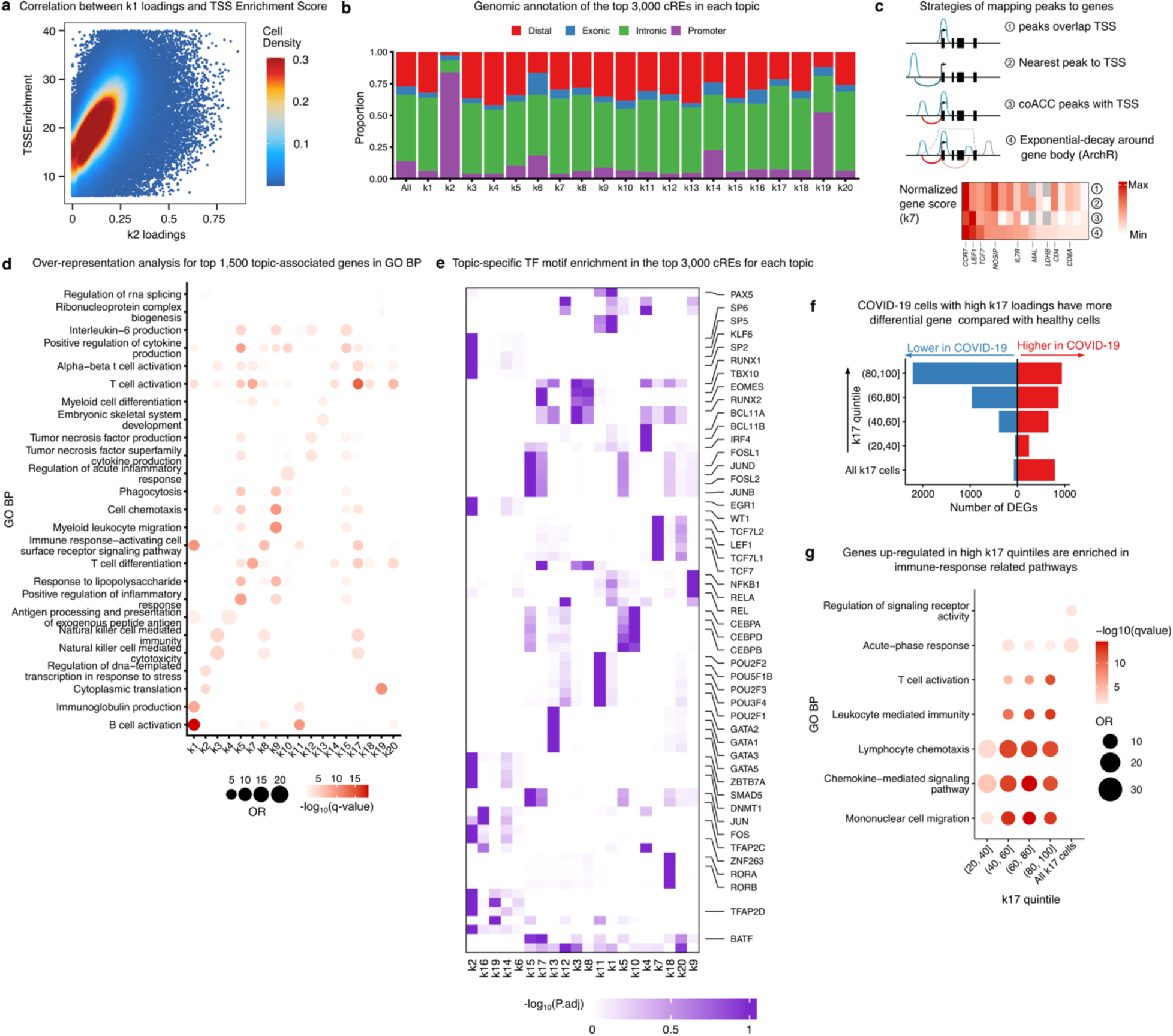
Topic modeling analysis. a, Scatter plot showing the correlation between k2 loadings and TSS Enrichment scores. **b**, Bar plot showing the genomic annotation of the top 3,000 peaks in each topic, compared to all peaks, highlighting the over-representation of promoters in k2. **c**, Top: schematic showing the four strategies to calculate gene-level score from peak-level scores in topic analysis. Bottom: gene-level scores calculated from the four strategies for peaks in topic k7, which is a naive T cell topic. **d**, GO Biological Process pathways enriched in top 1,500 scored genes for each topic. **e**, Heatmap of adjusted p-values for TF motif enrichment in top 3,000 peaks in each topic. -log10(P adj.) values are normalized relative to maximum for each TF across all topics. Top five enriched TFs are shown for each topic. **f**, The number of differentially active genes in COVID-19 cells in the top 4 k17 quintiles compared to all cells in the first k17 quintiles, plotted together with the number of differentially active genes when all COVID-19 cells in k17 were tested against all healthy cells in k17. **g**, GO Biological Process pathways enriched in up-regulated genes from groups in **f**. Only groups with significantly enriched GO terms were plotted.

**Extended Data Fig. 4.**
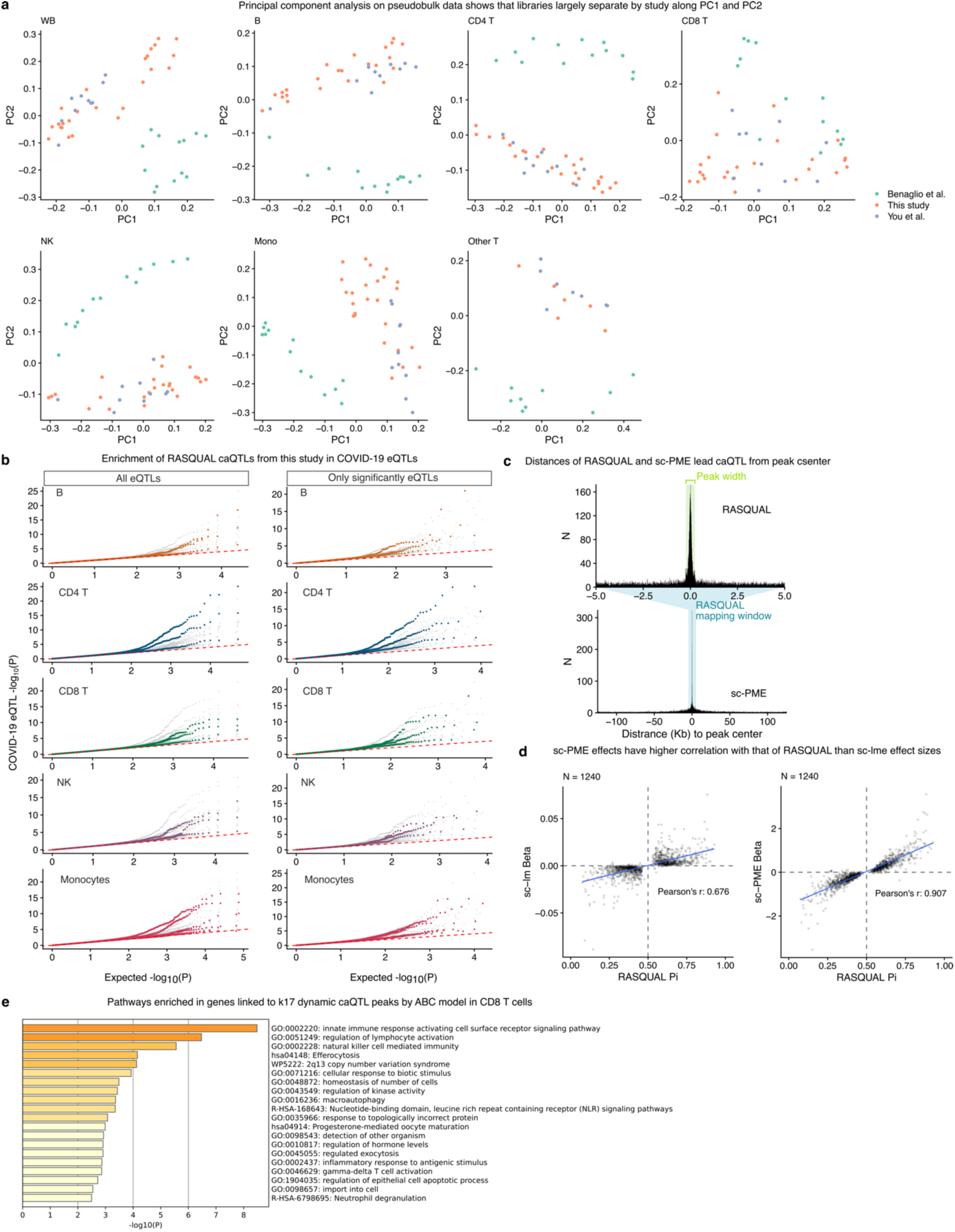
CaQTL mapping using RASQUAL and sc-PME model in harmonized data. a, Principal component analysis (PCA) on pseudobulk count data for WB and six cell-types in which caQTL mapping was conducted. Each sample is colored by study. **b**, QQ-plot showing the enrichment of caQTLs in our accompanying COVID-19 eQTLs for all eQTLs (left) and conditioning on only significant eQTLs (right). For each cell type in our study, we extracted and plotted eQTLs p-values from all cell types, highlighting matched cell types in colored dots; eQTL p-values from the remaining cell types were colored grey. When all genes are used, caQTLs tend not to have the highest enrichment in eQTLs in corresponding cell types, except for CD4 T and monocyte, due to lower power in the eQTL data (left). We therefore conditioned on only significant eGenes in each cell type to mitigate the differences in power between cell types, and observed larger enrichment of caQTL in eQTLs for matched cell types (right). **c**, Histogram showing distances from peak centers to significant lead caQTL in monocytes from RASQUAL (top) and sc-PME (bottom). Green shaded region highlights peak size (500 bp); blue shaded region highlights RASQUAL mapping window (10 Kb) relative to sc-PME mapping window (250 Kb). **d**, Scatter plots comparing RASQUAL effect sizes (Pi) with sc-lme effect sizes (left) and sc-PME effect sizes (right). Only significant RASQUAL caQTLs on chromosome 1 in monocytes were plotted. **e**, Top pathways enriched in genes linked to peaks with k17-interacting dynamic caQTLs in CD8 T cells.

**Extended Data Fig. 5.**
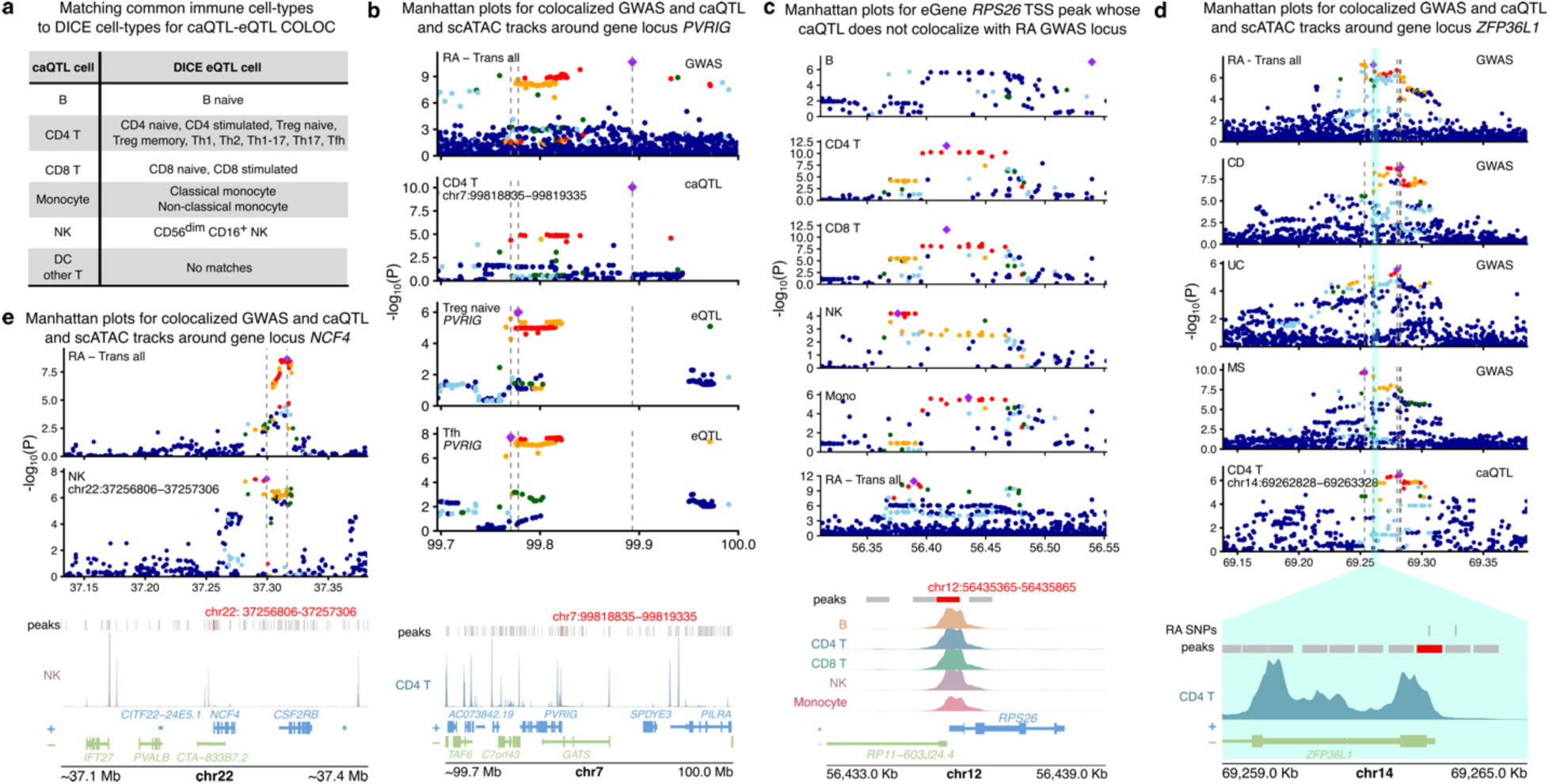
**a**, Table summarizing the mapping of 15 immune cell-types in DICE data to the seven common cell-types used in caQTL mapping. **b,** Manhattan plots for RA GWAS locus (7:99893148) and colocalized caQTLs (chr7:99818835−99819335) and eQTLs (*PVRIG*), with genome browser tracks for scATAC data in CD4 T cells. **c**, Manhattan plots for an RA GWAS locus near TSS of *RPS26*; the GWAS locus colocalized with *RPS26* eQTLs in DICE. The TSS peak is accessible and has caQTL in all five cell types, but the caQTLs do not colocalize with the RA GWAS. **d**, Manhattan plots for RA, CD, UC, MS GWAS loci and colocalized cPeak (chr14:69262828−69263328). Genome browser track shows promoter region of *ZFP36L1* near the colocalized peak and scATAC data in CD4 T cells. **e**, Manhattan plots for RA GWAS locus (22:37316259) and colocalized cPeak (chr22:37256806-37257306) in NK cells in the NCF4 locus. Genome browser track shows scATAC data in NK cells in the same region.

**Extended Data Fig. 6.**
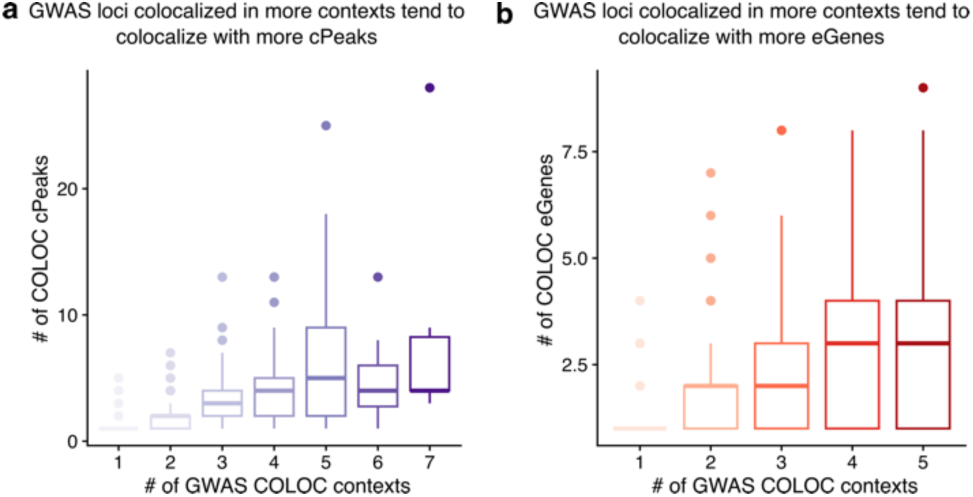
**a**, Boxplot showing the number of colocalized peaks for each GWAS locus as a function of the number of contexts where colocalization is detected. **b,** Boxplot showing the number of colocalized genes in DICE for each GWAS locus as a function of the number of contexts where colocalization is detected. The 15 DICE cell-types and subtypes were mapped to 5 common immune cell-types.

